# Insights into Renal Protein Handling Through GWAS of the Human Urine Proteome

**DOI:** 10.64898/2025.12.08.25341835

**Authors:** Stefan Haug, Oleg Borisov, Nick Lehner, Mona Schoberth, Irene Menneking, Katherine Xu, Atlas Khan, Nora Scherer, Sara Monteiro-Martins, Burulca Uluvar, Werner Römisch-Margl, Matthias Arnold, Pascal Schlosser, Peggy Sekula, Florian Kronenberg, Kai-Uwe Eckardt, Krzysztof Kiryluk, Matthias Wuttke, Gabi Kastenmüller, Yong Li, Anna Köttgen

## Abstract

The kidneys are an important determinant of circulating protein levels that retain plasma proteins at the glomerular filter and reclaim filtered proteins from urine. While genetic determinants of the circulating proteome have been extensively studied, studies of the urine proteome are lacking. We performed genome-wide association studies (GWAS) of 9 million common genetic variants for each of 2,868 proteins quantified using antibody-based technology from the urine of 1,246 persons.

We identified associations between urine protein levels and at least one significant (P<1.7E-11) variant at 174 loci. Index variants were mostly (153/174) located near their encoding *cis*-gene and enriched for protein-altering variants. Comparative analysis with plasma levels of the respective protein revealed a shared genetic basis at 122 loci, suggesting that their levels in urine provide a readout for systemic processes. Conversely, 38 loci showed urine-specific genetic signals. Variants in *CUBN* and *LRP2*, encoding the major receptor complex for tubular protein reabsorption from renal ultrafiltrate, were associated with >20 urine proteins each and shared with urine albumin, a known CUBN ligand. Integration with molecular and clinical data suggested many functional relationships, linking genetically driven changes in urine protein levels to differential gene expression, metabolite levels, and/or risk of disease. For example, genotypes at variants in the *PSCA* locus were perfectly predicted by urine PSCA levels and associated with *PSCA* expression and diseases of PSCA-expressing tissues, including bladder cancer and urinary tract infections, demonstrating potential clinical utility of urine protein levels.

These findings are accessible through a convenient web application, and establish a comprehensive genetic framework for kidney protein handling to facilitate clinical and experimental investigations.

## Introduction

The kidneys continuously clear plasma from waste products while retaining valuable solutes, thereby maintaining body homeostasis. This occurs via a complex process of glomerular filtration followed by substrate-specific tubular secretion and reabsorption processes, resulting in the precise control of the excreted amount of filtered solutes. Compared to plasma, urine is often considered “protein-free”, although it is known to contain small amounts of thousands of proteins^1^. Many of these proteins originate from plasma, as low-molecular-weight proteins pass the glomerular filter relatively freely^2^. Healthy individuals excrete up to 150 mg of protein per day^3^. Increased levels of proteinuria are not only used to diagnose and stage chronic kidney disease (CKD), but have also been identified as a strong risk factor for CKD progression^4^. Consequently, it is important to understand the mechanisms shaping the identity and amount of proteins present in urine, and how these may be modulated.

The kidneys have highly efficient tubular reabsorption mechanisms to prevent the loss of filtered proteins. The best-known mechanism of tubular protein reuptake is via the cubilin-megalin complex, which facilitates reabsorption of albumin, vitamin-binding proteins, enzymes, hormones, and others through receptor-mediated endocytosis^5^. Patients with biallelic mutations in the encoding genes, *CUBN* and *LRP2*, typically show low-molecular proteinuria^6,7^, and have informed the identification of ligands of the receptor complex. For instance, patients with Imerslund-Gräsbeck syndrome resulting from rare *CUBN* mutations^8^ suffer from vitamin B12 deficiency, enabling the identification of intrinsic factor-vitamin B12 as a ligand.

Another source of urine proteins is the cells lining the nephron and the urinary tract, from where they can originate through active secretion or shed cells. Secreted proteins include some with important roles in pathogen defense, such as uromodulin, which protects against urinary tract infections by *E. coli*^9^ and kidney stone formation^10^. Uromodulin and another tubular protein, EGF, indicate tubular health and integrity. Conversely, urine can also contain proteins that indicate the presence of pathogenic states such as tubular damage (e.g., KIM-1, LCN2), immune-mediated kidney diseases (e.g., complement proteins), as well as immune-related molecules in the presence of kidney cancer^11^.

Despite the kidney’s clearly important role in maintaining urine as well as circulating protein levels^12,13^, the mechanisms by which it handles most individual proteins remain unknown. Forward genetic mapping studies are a powerful way to identify genes that influence individual protein levels and mechanisms shaping the human proteome. However, published studies have almost exclusively focused on the circulating proteome^14–20^. We have previously shown for the human metabolome that 40% of findings identified in genetic association studies of metabolite levels are only detected when studying urine but not when studying plasma, and that such findings can reveal kidney-specific processes^21^. We therefore hypothesized that genetic studies of the urine proteome provide insights into mechanisms maintaining circulating and excreted protein levels beyond those enabled by the study of the plasma proteome. Here, we performed genome-wide association studies (GWAS) of the human urine proteome to identify genetic variants influencing the levels of nearly 3,000 urinary proteins to facilitate a better understanding of renal protein handling in humans and to identify specific urine proteins with the potential as biomarkers in an easily accessible fluid.

## Results

We conducted genome-wide studies to detect associations between 9,267,848 genetic variants of minor allele frequency (MAF) >1% and the levels of 2,886 protein analytes capturing 2,868 unique proteins from urine samples (**Figure 1 and Supplementary Figure 1)**. Samples were collected from 1,246 European-ancestry participants of the German Chronic Kidney Disease (GCKD) study^22^ (mean age 62 years, 60% men, median urinary albumin-to-creatinine ratio 13.7 mg/g; **Supplementary Table 1**). Protein levels were measured by Olink antibody-based proximity extension assay technology and corrected for inter-individual differences in urine dilution (Methods; **Supplementary Table 2**). A total of 994 and 658 proteins were detectable above the limit of detection in at least 50% and 80% of the samples, respectively. Thus, our data support the presence of hundreds of proteins in the urine of most individuals.

**Figure 1:**
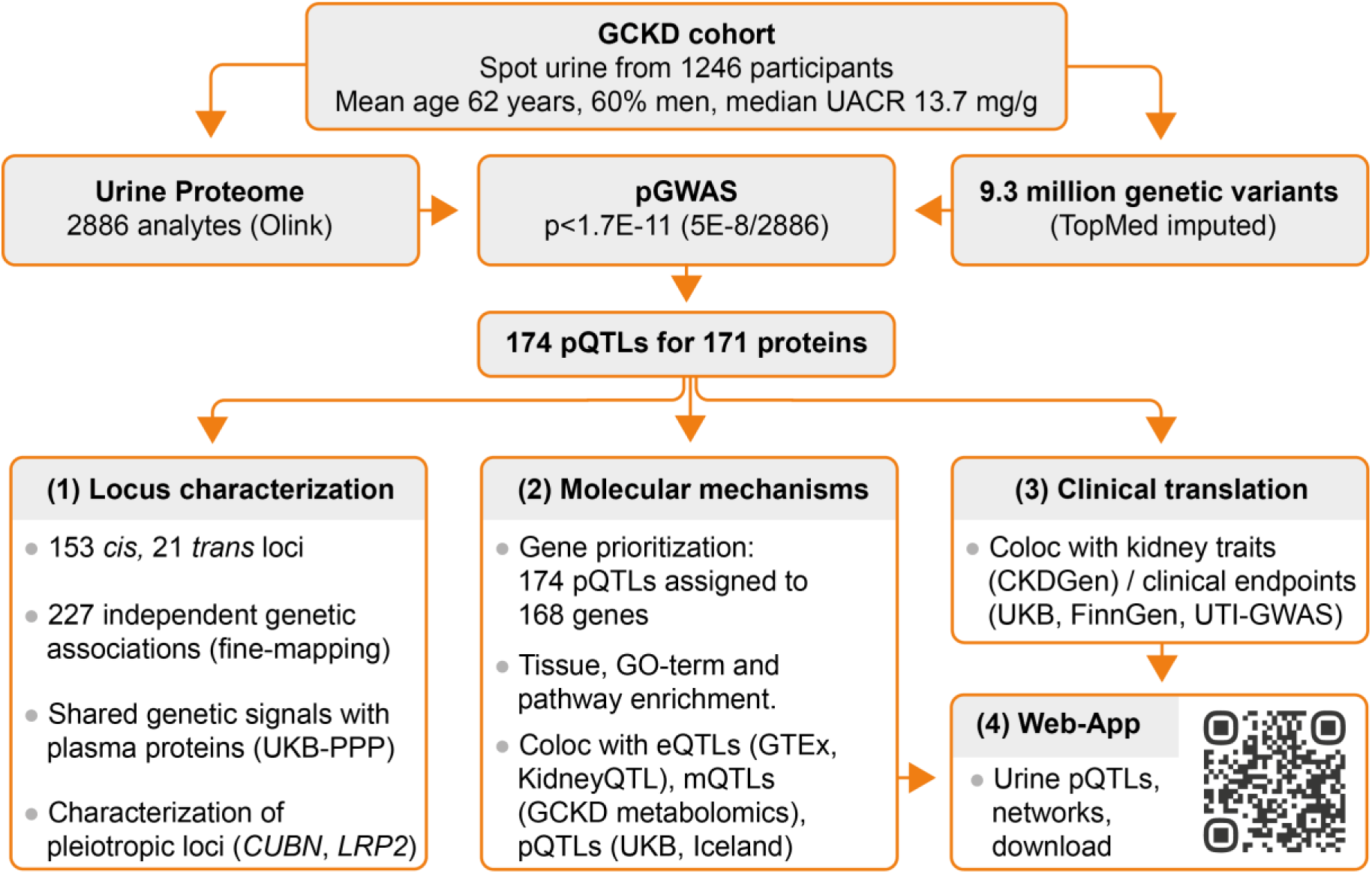
Overview of the study design. Schematic representation of genome-wide association studies for urine protein levels (pGWAS) and subsequent follow-up analyses. The interactive web application is accessible at: http://gwas.eu/urine-p/ (freely available after successful peer-reviewed publication). Abbreviations: GCKD, German Chronic Kidney Disease study; UACR, urine albumin-to-creatinine ratio; pQTL, protein quantitative trait locus; Coloc, genetic colocalization; UKB-PPP, UK Biobank Pharma Proteomics Project; GO, Gene Ontology; GTEx v8, Genotype-Tissue Expression project version 8; CKDGen, CKD Genetics Consortium, UTI, urinary tract infection.

### Identification and properties of 174 significant gene-protein associations

We detected 174 unique, covariate-adjusted protein-variant associations mapping to 127 distinct autosomal or X-chromosomal regions that contained at least one significant (P<1.73E-11; 5E-8/2886) variant. For each protein–region pair, the variant with the lowest association P-value was selected as the index variant and defined as a protein quantitative trait locus (pQTL; see Methods).

Across pQTLs, the explained variance of urine protein levels ranged from 3% to 67% (**Figure 2**). The associated protein was encoded in *cis* for 153 pQTLs (distance to the transcription start site of the protein-encoding gene ≤500 kb), whereas 21 pQTLs showed associations in *trans* (**Figure 2**, **Supplementary Table 3**). Three proteins, CD164L2, ICAM2, and SELPLG, had associated pQTLs in at least two loci. Across the urine proteome, the association with the lowest P-value (P=1.2E-451) was observed between a common (MAF 44%) missense variant in *PSCA* and urine levels of the encoded prostate stem cell antigen. The greatest effect size was observed for a *cis*-association between a common (MAF 8%) indel variant at *FOLR3* and levels of the encoded folate receptor gamma (1.7 standard deviations per copy of the minor CTA allele, P=1.1E-178; **Supplementary Table 3**). As expected, the genetic effect size was inversely related to MAF (**Supplementary Figure 2**).

**Figure 2:**
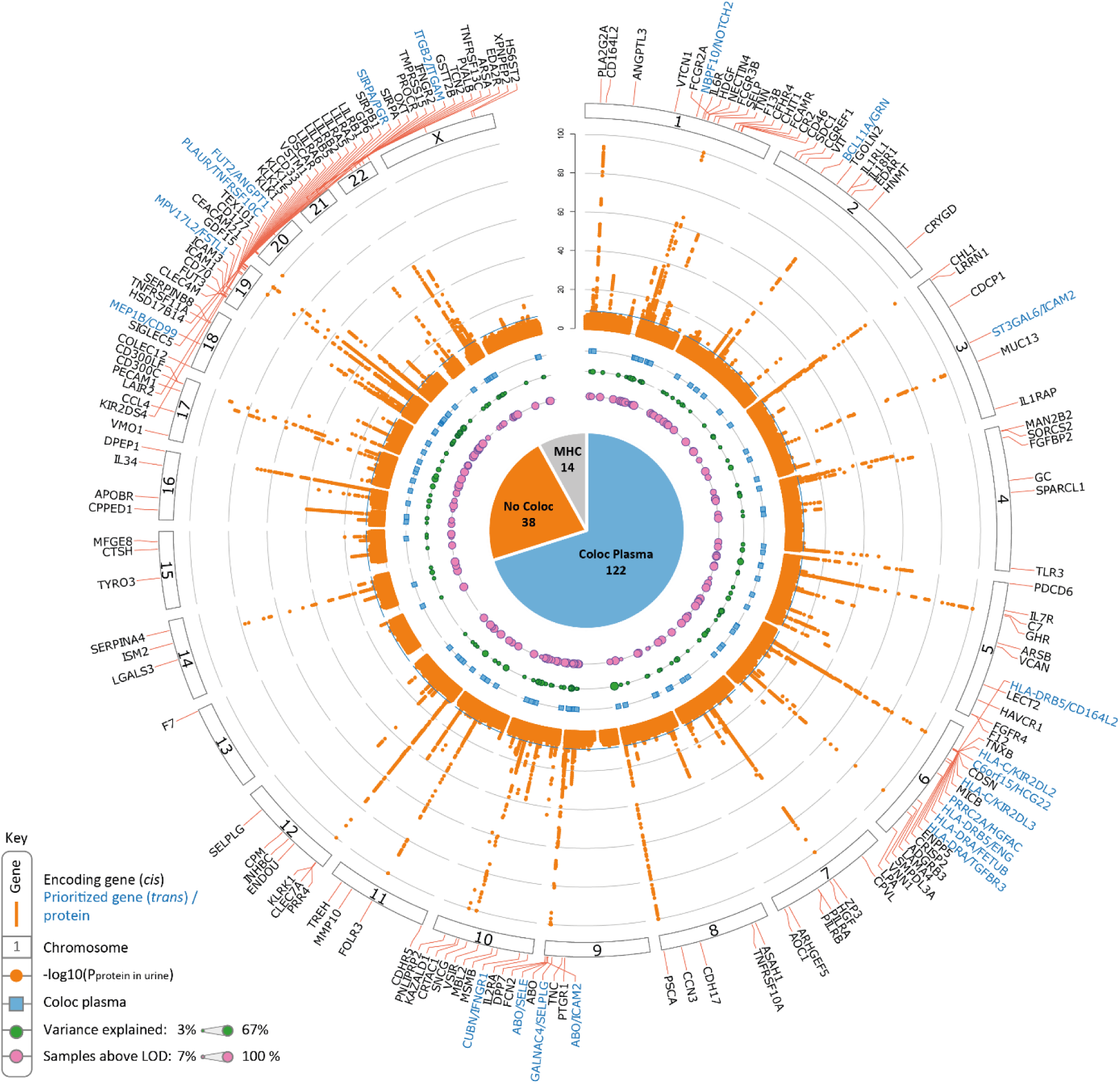
Circular plot of genetic associations with urine proteins and overlap with respective plasma proteins. Circos plot summarizing genetic associations with levels of 2868 urinary proteins. The orange dots correspond to the –log10(P-value) of the association between a genetic variant and urine protein levels and are ordered by chromosomal position, with the y-axis truncated at 100. The horizontal blue line indicates study-wide significance (P=1.7E-11). Supplementary Table 3 provides details on index SNP associations (pQTLs). *Cis*-pQTLs were assigned to genes encoding the associated protein (black labels); *trans*-pQTLs (blue labels) were assigned to prioritized genes based on proximity, pQTL annotations and colocalization analysis with gene expression and protein levels (Methods). The pie chart displays the proportion of pQTLs with and without colocalizing genetic associations with plasma levels of the respective protein (PP H4 > 0.8). The 14 pQTLs labeled with “MHC” also include three pQTLs for which a colocalization analysis was not possible due to a low number of overlapping variants in the plasma data. The inner rings display three data types: blue squares indicate the 122 loci with positive plasma colocalization, green circles show the variance in urine protein levels explained by each pQTL, and pink circles represent the percentage of samples with detectable protein levels above the limit of detection (LOD) for pQTL-associated proteins.

The most likely causal gene at each locus was assigned as the protein-encoding gene for *cis*-associations. For *trans*-associations, a workflow that considered proximity, variant annotations, as well as colocalization with gene expression and protein levels was implemented to assign the most likely effector genes (Methods; **Supplementary Table 3**). Several analyses were conducted to detect biological pathways, molecular functions, cellular components, as well as tissues and cell types in which prioritized genes were over-represented (Methods). Prioritized genes were predominantly enriched in terms related to the extracellular region/space and cell periphery, to plasma membrane, and to (transmembrane) immune-mediated signaling after correction for multiple testing (FDR P-value<0.01) and using Olink panel genes as background (**Supplementary Table 4**). When using all protein-coding genes as background, additional terms related to immune-mediated signaling showed strong associations, possibly influenced by Olink assay design (**Supplementary Table 5**). Prioritized genes showed enhanced transcript levels in liver, immune-related tissues, lung, intestine, and kidney (**Figure 3a, Supplementary Table 6**). Cellular compartments in which prioritized genes were significantly enriched at the protein level (IHC-based) were microvilli of gut enterocytes, microvilli of kidney proximal tubular cells (driven by DPEP1, PROCR, GHR, TREH, XPNPEP2, MFGE8, and FCAMR), as well as smooth muscle cells of the endometrium (**Figure 3b, Supplementary Table 6**).

**Figure 3:**
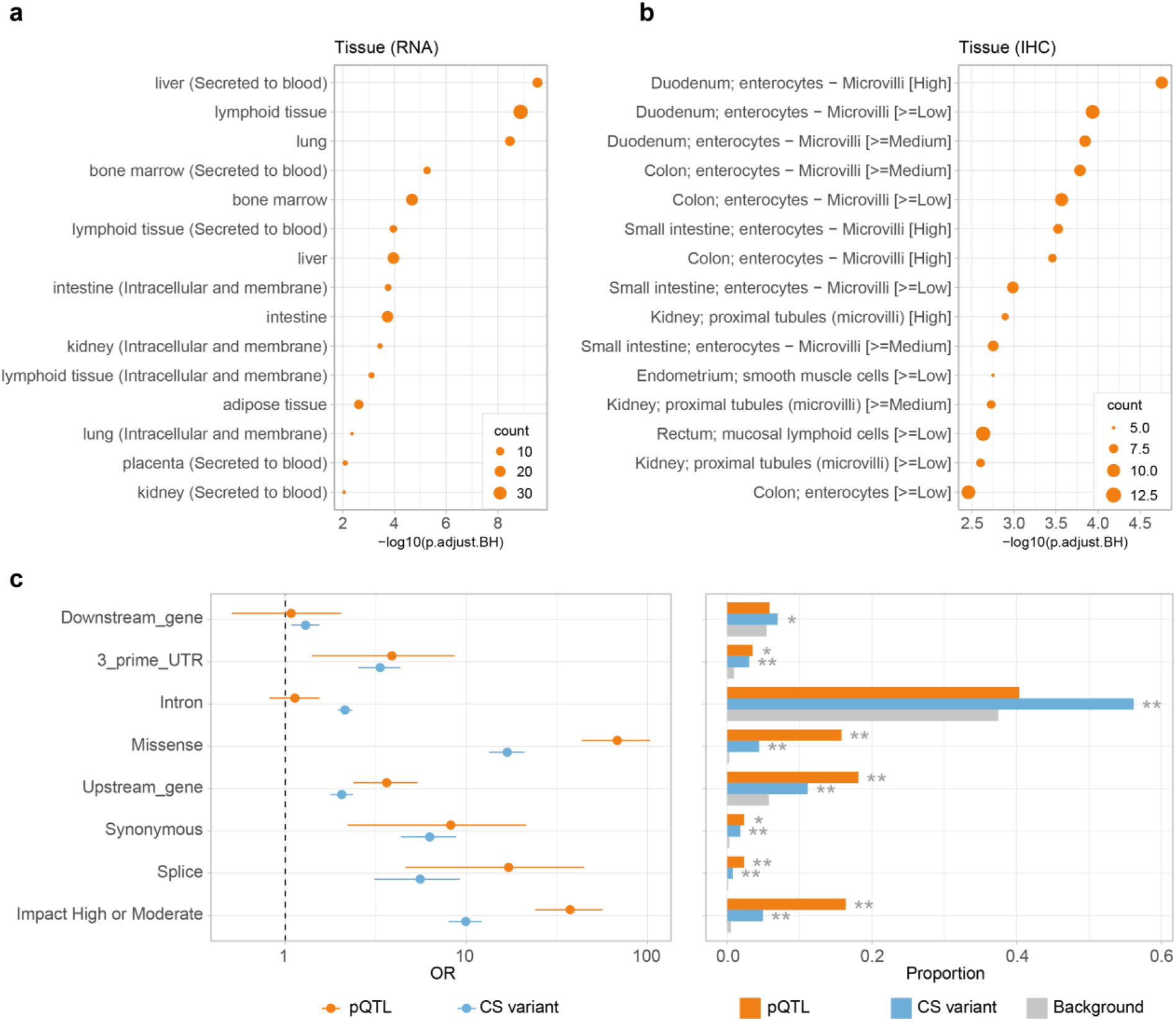
Tissues enriched for enhanced expression of pQTL effector genes and enriched pQTL variant consequences. **a**, Tissue enrichment analysis for prioritized genes from all pQTLs. Each tissue was defined by genes with enhanced expression in that tissue. Tissue subcategories were created by further stratifying these tissue-enriched genes based on their secretory status (e.g., “secreted to blood“ or “intracellular and membrane“). **b,** Enrichment analysis of prioritized pQTL genes across cellular compartments at the protein level. Genes were assigned to cellular compartments based on immunohistochemistry (IHC) expression grades from the Human Protein Atlas (≥low, medium, or high intensity). **c**, Enrichment analysis for pQTL variant annotation categories. The left panel shows odds ratios (OR) and 95% confidence intervals (horizontal bars) from Fisher’s exact test comparing variant annotation categories between: (i) 171 unique pQTLs (orange), (ii) 1,943 unique credible set (CS) variants with conditional *P*<5E-8 and PIP≥0.01 (light blue), and (iii) all variants tested in this association study (background, gray). The dotted line marks OR = 1. The right panel shows the proportion of variants in each annotation category for pQTLs, CS variants, and background variants. Fisher’s test: *P*<0.05, \*\**P*<0.001. Only annotation categories with more than two annotated pQTLs are shown.

### Fine-mapping and annotation of candidate causal variants

Statistical fine-mapping indicated the presence of 227 credible sets within 163 associations outside the extended Major Histocompatibility Complex (MHC) region (range 1-8 per locus; Methods; **Supplementary Table 7**). Forty-seven credible sets contained a single variant causing the observed association signal with >95% posterior probability (PP), and 107 credible sets had 5 or fewer variants. The median PP across the 156 pQTLs contained in any credible set was 31% (Q1: 13%, Q3: 67), as compared to 4% for all variants across credible sets (**Supplementary Table 8)**. The 171 unique pQTLs had a significantly higher chance of being annotated as missense, high or moderate impact, splice, and other functional consequences compared to background variants (**Figure 3c**). For example, pQTLs had a 68 times larger chance of being annotated as missense compared to the background (**Supplementary Table 9**). **Supplementary Table 8** contains detailed annotation of credible set variants based on several kidney-specific functional genomics datasets (Methods) as an informative resource to prioritize not only protein-altering, but also potentially regulatory variants for experimental follow-up studies.

### Comparative analyses with plasma pQTLs highlight urine-specific and shared signals

Genetic associations with urine protein levels that are also detected in plasma could, especially for low-molecular weight proteins, result from their systemic production followed by glomerular filtration. Conversely, different association signals in the same region or associations with protein levels solely detected in urine may contain information about kidney-specific processes. We performed a large-scale colocalization screen for the 163 urine pQTL regions outside the MHC to assess whether they share a genetic basis with plasma protein levels, using published GWAS of plasma levels of 2,923 proteins from 34,090 EUR participants of the UK Biobank^20^, as well as 4,907 protein-capturing aptamers from 35,559 Icelanders^18^ (Methods; **Supplementary Table 10**). We detected colocalization (PP H4>0.8; Methods) that involved at least one plasma protein with a genome-wide significant (P<5E08) association signal in 2,498 instances. The majority of the colocalizing plasma proteins (2,342) were encoded in *trans*. There were 742 colocalizing plasma proteins quantified by Somalogic technology and 1,756 by the same Olink assay as used for urine, with 267 of them identified by both proteomics technologies. Moreover, we identified 26 instances in which levels of two urine proteins shared a genetic origin.

Since the colocalization screen revealed substantial overlap between the genetic architecture of urine and plasma protein levels, we next asked how many urine pQTL regions shared an association signal with the same protein in plasma. We therefore performed fine-mapping analyses using individual-level data from our urine study and the UK Biobank plasma proteomics study to account for the potential presence of multiple independent signals per region, followed by pairwise urine vs. plasma colocalization of all identified signals within each pQTL region (Methods).

At 122 of 163 urine pQTL regions outside the MHC locus and with sufficient genetic coverage, we identified at least one credible set of SNPs that shared associations with the levels of the respective protein in plasma (**Figure 2**). Conversely, 38 loci did not show evidence of a shared genetic basis for protein levels in urine and plasma (**Supplementary Table 11**, **Supplementary Data 1**). Five of these loci did not contain any genome-wide significant (P<5E-08) genetic variant associated with the levels of the implicated protein in plasma despite a 27-fold larger sample size of the plasma screen, namely at prioritized genes *BCL11A*, *ISM2*, *FUT2*, *MPV17L2*, and *CUBN* (further discussed below). The remaining 33 loci showed evidence for different variants in the locus giving rise to associations with plasma and urine protein levels, such as at *VTCN1*, *LAMA4*, *CPVL*, *AOC1*, or *TMPRSS15*. Such urine-specific association signatures might implicate kidney cell type-specific regulatory variants, detailed functional annotation of which including kidney cell type-specific chromosomal accessibility is provided in **Supplementary Table 8.**

### Integration with gene expression and metabolomics: molecular mechanisms

We also used genetic colocalization testing to systematically integrate urine pQTL regions with levels of gene expression, metabolites, or urine protein levels with at least one suggestive association signal (P<1E-05; Methods). The results are publicly accessible through a web application (http://gwas.eu/urine-p/ (freely available after successful peer-reviewed publication)) to facilitate visualization and exploration of hypotheses about potential molecular mechanisms. A noteworthy example of the integration across urine proteins (**Supplementary Table 10**) was detected at the *CUBN* locus, where the same genetic variants were driving the association not only with urine levels of interferon gamma receptor 1 (IFNGR1), but also of 25 additional proteins (**Figure 4**), of which 23 were affected in the same direction as urine IFNGR1 levels.

**Figure 4:**
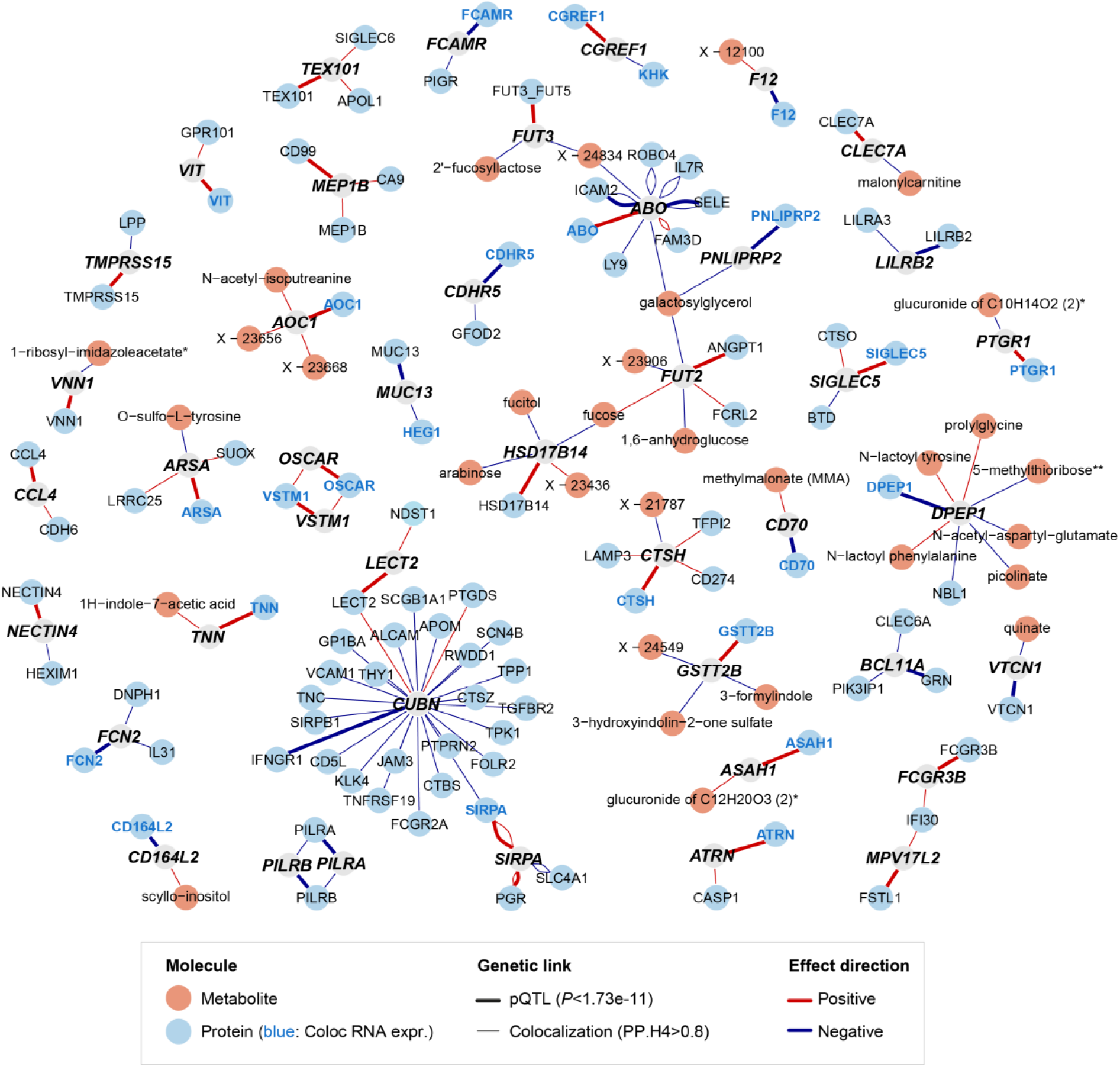
Genetic colocalization network of urine proteins and metabolites. Network representation of pQTLs with at least one colocalizing association with a urine metabolite or another urine protein. pQTLs are shown as gray circles, labeled with their prioritized effector gene (**Supplementary Table 3**), and linked to the associated protein with a bold line (multiple lines for pleiotropic loci). Thin lines indicate urine metabolites or other urine proteins with colocalizing genetic signals. All associations with urine proteins are derived from this study; metabolite associations from the same study sample were published previously^41^. The pQTL-associated protein is highlighted in blue if colocalization with the encoding transcript was detected in any GTEx v8 tissue. Effect directions of the pQTL index variant (effect allele according to **Supplementary Table 3**) on urine levels of the associated proteins, colocalizing proteins, and colocalizing metabolites are color-coded as red (positive) and blue (negative). An extended interactive colocalization network including additional association data with clinical traits (FinnGen, UK Biobank, CKDGen Consortium) and plasma proteins (UK Biobank, Icelanders) can be accessed through our web application: https://metabolomips.org/koettgenpgwas/ (Username: kgwas, Passcode: web).

Tissue transcriptomics provided evidence for a shared genetic basis of urine protein levels and gene expression for multiple plausible examples (**Supplementary Table 12**). There were several regions in which the same genetic variant was driving the association with gene expression in kidney tissue and the levels of a urine protein encoded in *cis*, such as at *DPEP1*, *ABO,* and *GSTT2B* (**Figure 4**), encoding enzymes expressed in kidney tissue. While these observations support that urine protein levels may reflect the differential regulation of their cognate genes’ expression in the kidney, exploring extra-renal tissues provided additional insights. For instance, genetic variants in the *ITGB2* locus, encoding integrin beta 2, were associated with *ITGB2* transcript levels in many tissues and with the levels of integrin alpha M (ITGAM) protein in urine. Although ITGAM is encoded on a different chromosome, it forms a leukocyte-adhesion-related heterodimer with integrin beta 2, linking the genetic association to a functional protein complex.

Further integration of circulating and urine metabolite levels (**Supplementary Table 13**) provided additional, complementary information, often supported by existing biological knowledge. For instance, the same variants associated with urine protein levels of glutathione S-transferase theta 2B (GSTT2B), an enzyme with sulfatase activity^23^, were associated with the levels of 3-hydroxyindolin-2-one sulfate in urine (**Figure 4**), as well as with several other indole-containing metabolites in plasma and urine. Similarly, protein levels of glycosyltransferases ABO and FUT2 in urine showed colocalization with the urine levels of galactosylglycerol, levels of the kidney enzyme amine oxidase 1 (AOC1) with the metabolite putreamine in urine, and levels of renal dipeptidase 1 protein with the levels of several dipeptide metabolites in urine. The A allele at the *DPEP1* index variant, rs2434858, was associated not only with lower levels of renal *DPEP1* expression and lower urine DPEP1 levels, but also with higher levels of prolylglcyine in urine, consistent with its role as a dipeptidase that may hydrolyze this metabolite (**Figure 4**, webserver). Lastly, we found a shared genetic basis of the urine levels of HSD17B14 and fucose, supporting the recent detection of hydroxysteroid 17-beta dehydrogenase 14 (HSD17B14) as the long-sought mammalian L-fucose dehydrogenase^24^ (**Supplementary Figure 3**), and highlighting the potential of our data to explore new hypotheses via convenient webserver access.

### The cubilin/megalin receptor complex shapes the urine proteome

The kidneys contain an efficient system for the reabsorption of filtered proteins to prevent their loss. The major complex for receptor-mediated protein endocytosis consists of cubilin and megalin, two large, multiligand glycoproteins^25–27^. We observed a urine-exclusive *trans*-association between common variants in the *CUBN* locus, encoding cubilin, and urine levels of interferon gamma receptor 1 (IFNGR1; P=3.9E-15). The intronic index variant was in high LD (r^2^=0.97) with the missense variant rs62619939, encoding p.Leu2153Phe in cubilin (ENSP00000367064.4; **Supplementary Table 8**). Interestingly, the shared genetic basis between IFNGR1-associated *CUBN* variants and 25 additional urine proteins was also observed for one plasma protein, amnionless (AMN; **Supplementary Table 10**). AMN is a type I transmembrane protein, which, together with cubilin, forms part of the cubilin-megalin receptor complex. Rare mutations in *CUBN*, including p.Leu2153Phe^8^, as well as in *AMN* can both cause Imerslund-Gräsbeck syndrome, an autosomal-recessive condition featuring vitamin B12 deficiency and proteinuria^6,28^. These observations are extremely unlikely to arise by chance. Together with the identification of colocalizing urine proteins that are known cubilin ligands, such as Clara cell secretory protein (SCGB1A1)^29^, they support the biological plausibility of our findings and accurate protein targeting.

We next explored the existence of additional pleiotropic regions for urine pQTLs of at least genome-wide significance (P<5E-08) by searching for shared genetic determinants with the levels of other urine proteins (Methods). Across the genome, we identified three loci at which a shared variant was underlying associations with at least 20 proteins, mapping into *CUBN*, *LRP2*, and an intergenic region on chromosome 4 (**Figure 5a, Supplementary Table 14**). The detection of *LRP2*, encoding the cubilin-complex member megalin, out of approximately 20,000 protein-coding genes is highly unlikely to occur by chance, and supports that common variants in the genes encoding this receptor complex shape the human urine proteome. The expression of the complex at the urine-facing apical membrane of tubular epithelial cells is consistent with the total lack of associations with plasma protein levels.

**Figure 5:**
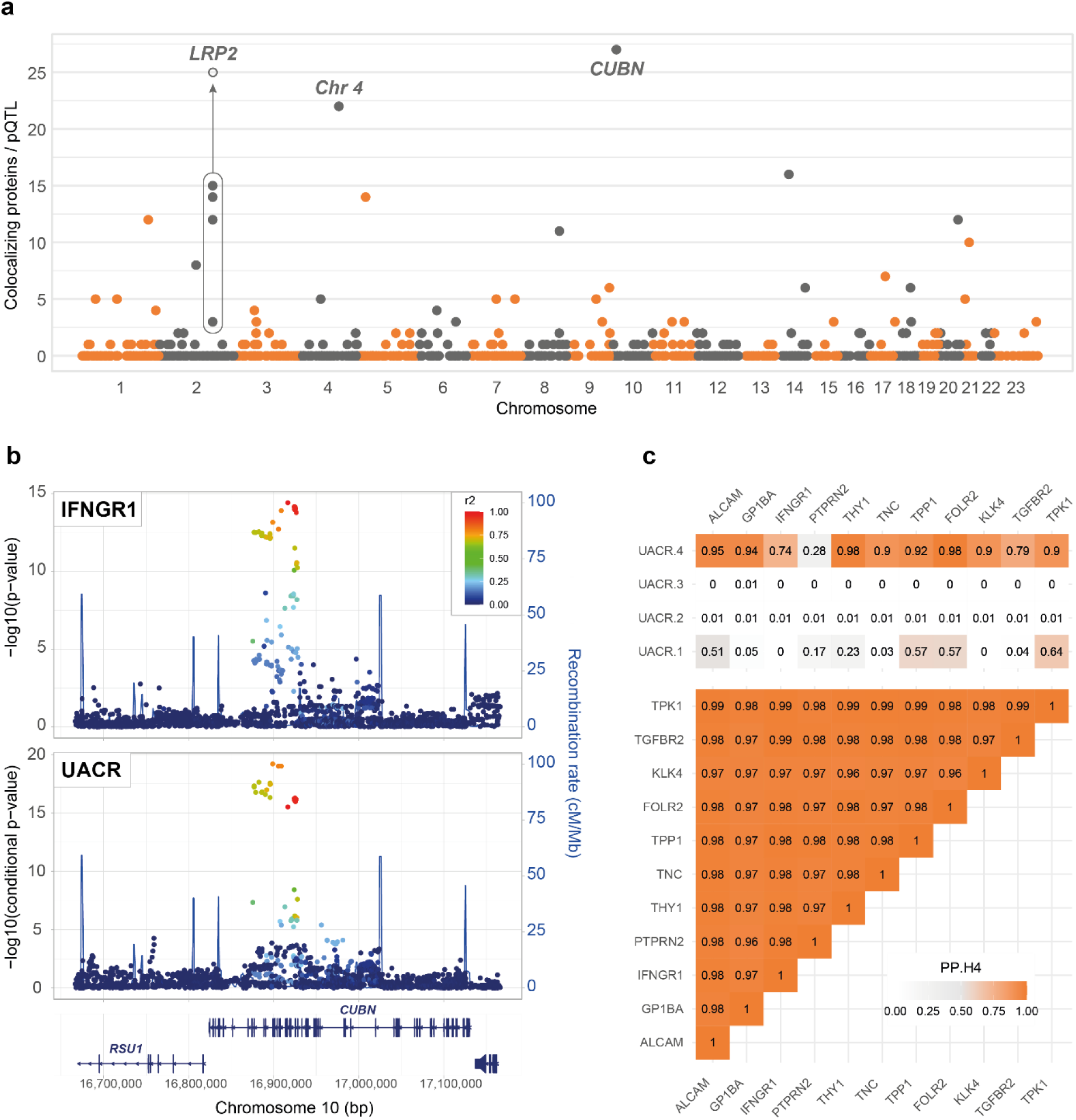
*CUBN* and *LRP2* are pleiotropic loci that shape the urine proteome. **a**, Colocalization-based screen for pleiotropic urine protein-associated loci. The dots indicate the number of colocalizing urine proteins per pQTL and are positioned at each pQTL’s chromosomal location. pQTLs for this pleiotropy screen were identified with *P*<5E-8 (*N*=958), and genetic colocalization was performed for all urine proteins with at least suggestive associations (*P*<1E−5) in each pQTL region. Three loci show more than 20 colocalizing distinct urine proteins: *CUBN*, *LRP2*, and an intergenic region on chromosome 4. The *LRP2* locus has a complex genetic architecture with multiple pQTLs that collectively colocalize with 25 urine proteins, as indicated by the gray circle. **b**, Regional association plots for the *CUBN* locus showing the association with urine IFNGR1 levels (top) and UACR (bottom, UK Biobank data). Conditional *P*-values are shown for UACR, and the independent signal corresponds to UACR.4 in panel c. **c**, Genetic colocalization between UACR and urine levels of multiple proteins at the *CUBN* locus. The top panel illustrates the PP.H4 values between each of four conditionally independent UACR signals and all urinary proteins with association *P*<1E-6. The protein levels show near-perfect genetic colocalization among each other (bottom panel).

Common genetic variants in the *CUBN* locus were previously shown to associate with levels of the urine albumin-to-creatinine ratio (UACR) in large population studies^30^. Using data from the UKB^31^, we tested whether the same variants in the *CUBN* locus were responsible for the known association with UACR and the urine proteins detected in our study. Indeed, we detected colocalization between conditionally independent variants in the *CUBN* locus with UACR and with IFNGR1 levels (**Figure 5b, c**; PP H4=0.74), with regional association plots for the other colocalizing proteins shown in **Supplementary Figure 4**. The shared genetic architecture between UACR and urine protein levels further extended to the levels of ALCAM, GP1BA, THY1, TNC, TPP1, FOLR2, KLK4, TGFBR2, and TPK1 (**Figure 5c**; PP H4 0.79-0.98). Notably, the same allele was associated with higher UACR and with higher urine protein levels of all mentioned proteins. These findings implicate these proteins as potential cubilin and/or megalin ligands, a hypothesis further supported by the recent reporting of TPP1, THY1 and TNC as potential megalin/cubilin substrates from a kidney-specific Lrp2 knockdown mouse model^32^.

Lastly, the identified pleiotropic region on chromosome 4 deserves further study. While the locus did not contain any candidate gene obviously connected to protein endocytosis in the proximal tubule, it is noteworthy that two of the associated proteins, fatty acid-binding proteins 2 and 4, have recently been reported as potential megalin/cubilin ligands in a systematic screen using kidney-specific megalin knockdown mice^32^ .

### Urine levels of prostate stem cell antigen are a readout of disease-associated PSCA genotype

The *cis*-association with the lowest P-value in this study was observed between an intronic base insertion in *PSCA*, rs71514093, and urine levels of the encoded prostate stem cell antigen (**Supplementary Table 3**). Genotype at rs71514093 displayed a striking association with PSCA urine levels (**Figure 6a**), especially with dominant modeling of the guanine insertion (P-value=7E-506). Virtually all persons carrying at least one copy of the G insertion showed levels above Olink’s limit of detection for PSCA (**Figure 6a**), and urine PSCA levels were almost perfect predictors of carrier status of at least one G insertion at rs71514093 (area under the ROC curve = 0.99; **Figure 6b**).

**Figure 6:**
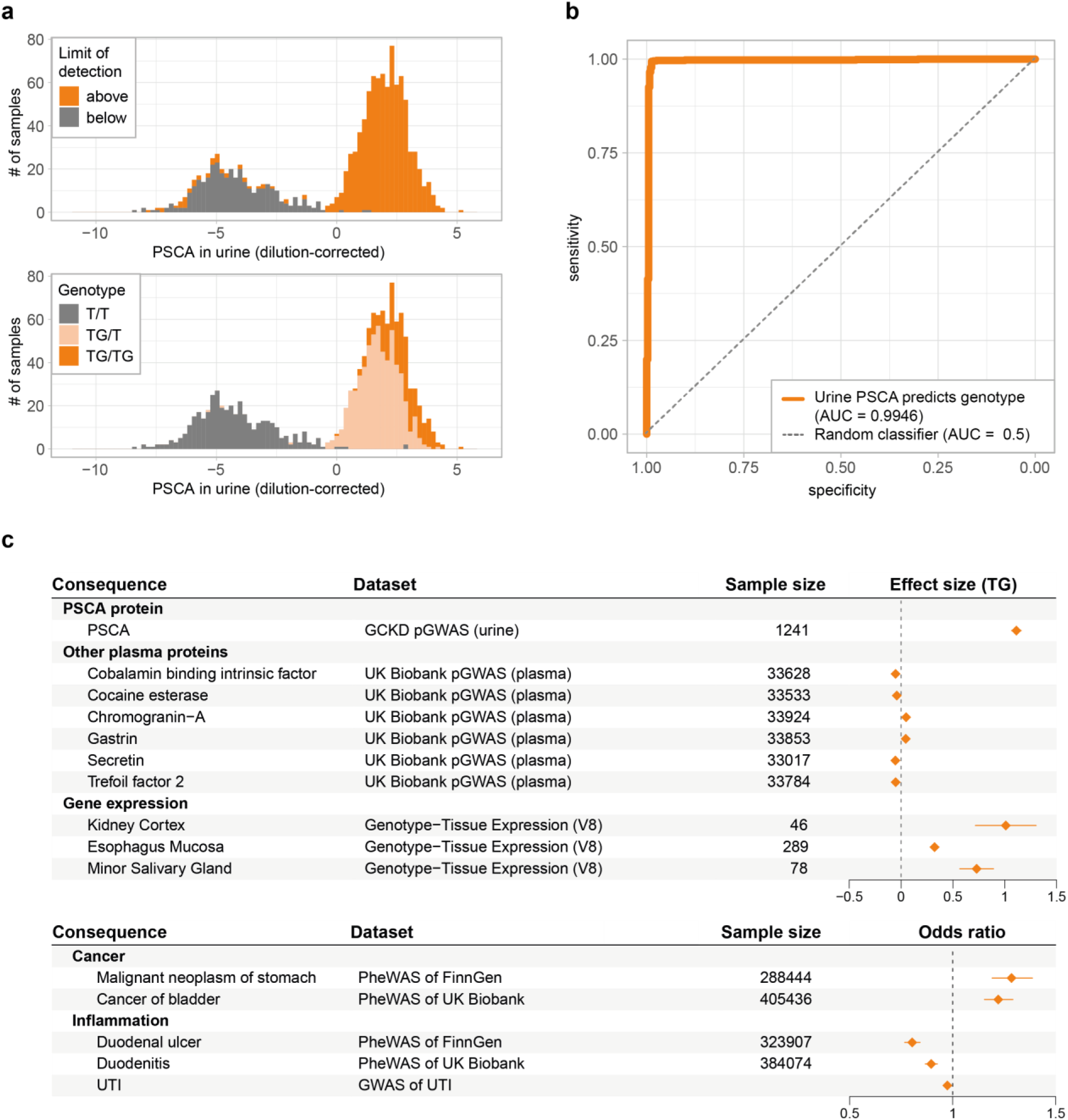
PSCA levels in urine predict disease-associated *PSCA* genotype. **a**, Distribution of dilution-corrected PSCA levels in urine, color-coded for samples with levels detected above and below the assay’s limit of detection (top) and for the urine donors’ rs71514093 genotypes (bottom). **b**, Receiver operating characteristic (ROC) curve for the prediction of rs71514093 genotypes (i.e. the presence of at least one TG allele) based on urine levels of PSCA. **c**, Association of rs71514093 with urine PSCA levels and other molecular and clinical traits. Traits were selected based on positive genetic colocalization (PP H4 >0.8) with the PSCA association signal in urine, indicating a shared causal variant. Effect size and odds ratio are shown for the TG allele. pGWAS: GWAS of protein levels; PheWAS: phenome-wide association study; UTI: urinary tract infection.

We detected a shared genetic basis of urine PSCA levels with the levels of circulating PSCA (**Supplementary Table 11**) as well as with *PSCA* transcript levels in 11 tissues, including several gastrointestinal tissues and the kidney (**Supplementary Table 12**; **Figure 6c**), supporting a regulatory effect of the underlying causal variant. The shared genetic basis extended to circulating levels of 10 additional proteins encoded in *trans*, three of which were supported by both proteomics platforms (**Supplementary Table 10**). Most of these 10 proteins have established roles in gastrointestinal physiology and digestion, including intrinsic factor, ghrelin, chromogranin-A, trefoil factor 2, and secretin. This mirrored the additional shared genetic basis between urine PSCA levels and 14 clinical traits and diseases (**Supplementary Table 15**), several of which were identified in different datasets and all pertained to gastroduodenal ulcers and stomach cancer on the one hand, and to bladder and kidney cancer and urinary tract infection (UTI) on the other hand. The G insertion at rs71514093 was associated with higher risk of bladder and stomach cancer, and with lower risk of duodenal ulcer and UTI (**Figure 6c**). These findings underscore the antagonistic pleiotropy between oncogenic processes and mucosal defense traits. Together, our data suggest that urine PSCA levels are an accurate readout of the PSCA-mediated genetic predisposition to cancers and infections of the urinary and gastrointestinal tract, where the protein is highly expressed (GTEx Project^33^ and Human Protein Atlas^34,35^).

## Discussion

This first study of the genetic basis of the human urine proteome has several key findings: first, almost a thousand proteins can be quantified above detection levels in the majority of spot urine samples, challenging the hypothesis that urine is largely protein-free and showing that urine is a biosample well suited for proteogenomic studies. Second, we detected 174 instances of significant associations between genetic variants and proteins even after stringent correction for multiple testing, highlighting genetic influences shaping the urine proteome. Third, while many of these associations can also be detected in plasma, there are urine-exclusive and urine-specific associations that cannot be identified when studying the circulating proteome. These include a footprint of the receptor-mediated protein endocytosis complex composed of cubilin and megalin on the urine proteome, including potential undescribed ligands of this complex. Fourth, as illustrated by PSCA, our findings show that urine proteins can serve as a readout of genotype and genetically-mediated risk for human diseases related to PSCA-expressing tissues, including diseases of the lower urinary tract. The strong urine cis-pQTL signal and its co-localization with GWAS for UTI nominates PSCA as an excreted molecule that is potentially protective against uropathogens, similar to the previously identified effect of uromodulin^9^. Further studies are needed to delineate the mechanism of this protective effect. Inverse risk relationships with higher risk of cancers and lower risk of autoimmune disease have previously been reported for genes encoding for immune checkpoint proteins^36^. Lastly, multi-Omics data integration, accompanied by a convenient web tool to interactively explore the results constitutes an important resource for the generation of hypotheses about disease-associated molecular mechanisms and serves as a starting point for experimental validation studies.

Previous genetic studies of the human proteome have almost exclusively focused on plasma^14,15,17–20^, which integrates information across dozens of tissues. However, a recent study of the cerebrospinal fluid proteome supports the notion that studies of biological fluids beyond plasma yield unique and complementary insights about the function of specific organs and related diseases^37^. In this first large-scale genetic study of the urine proteome, we show that these observations extend to urine, where urine-specific findings highlight processes known to specifically localize to the kidney such as the tubular protein reuptake machinery. This is in agreement with our previous studies of the urine as compared to the plasma metabolome, which showed that urine is a sensitive readout of metabolic processes happening at the apical membrane of tubular epithelial cells^21,38,39^.

A urine-exclusive finding of particular interest is the association between genetic variation in the *CUBN* and *LRP2* loci, encoding the two major components of the complex responsible for receptor-mediated protein endocytosis in the proximal tubule, and dozens of urine proteins. Endocytosis of filtered plasma proteins is essential to reclaim nutrients and to regulate hormone-mediated homeostatic processes^27^. Disruption of receptor-mediated endocytosis and its associated apical endo-lysosomal system causes proteinuria, a hallmark of kidney disease. Moreover, a better understanding of tubular protein reuptake may offer potential entry points for therapeutic targeting of proximal tubular cells. In a way, our study can be considered the human analogy to experimental studies that have aimed to identify ligands of the cubilin/megalin complex through targeted genetic manipulation^32^. Our study identified a shared genetic basis between common variants in the *CUBN* gene and the levels of urine albumin, a known ligand of cubilin, as well as with 25 other urine proteins, which may represent ligands of the receptor complex. In addition to some well-established ligands such as Clara cell secretory protein (SCGB1A1)^29^, they contain candidate ligands such as TPP1, THY1 and TNC that have recently been reported from a genetically manipulated animal model^32^. Thus, the ligand repertoire of this important mechanisms to reclaim proteins otherwise lost in the urine may be even larger than currently appreciated.

An important strength of our study is its unique design focused on the urine proteome, followed by a rigorous comparison to the circulating proteome quantified by the same technology. Since there were more than 27 times as many individuals in the genetic study of the plasma as compared to our study of the urine proteome, it can be expected that in studies of comparable sample size, even more loci would have been classified as urine-specific at a given level of significance. Another strength is the availability of urine and plasma metabolomics data from the same individuals, a complementary way to facilitate the detection of kidney-specific information. The efficient method for performing large-scale colocalization screens with thousands of molecular and clinical traits that we developed represents an important analytical resource. Potential limitations include the relatively small sample size of our study. However, genetic associations were robustly observed across two individual batches, and many were validated in the comparison to the circulating proteome. Although current proteomics platforms still have limited coverage as compared to the human transcriptome, we were able to compare all evaluated urine proteins to plasma by aligning proteomics platforms. The GCKD study is limited to European ancestry individuals by design, which may limit the generalizability of our findings. However, a previous study of the circulating proteome used for comparative purposes showed that genetic findings detected among European ancestry individuals were highly concordant with their effects in persons of non-European ancestry^20^. Most of the GCKD study participants have moderately reduced eGFR. However, our analyses adjusted for eGFR, and we have previously shown for metabolites that genetic effects on urine and plasma metabolite levels are comparable for GCKD study participants and persons with normal eGFR^21,38,39^. Moreover, shared genetic architecture between the identified urine pQTLs and the circulating proteome, quantified from persons with mostly normal kidney function, further support that are findings are largely unaffected by lower kidney function. Lastly, previous studies have shown that some plasma pQTL association are platform-specific, and that readouts may sometimes correlate with protein modifications or isoforms rather than their absolute levels^40^. It is therefore reassuring that for 47 of the urine pQTL loci, a shared genetic architecture was also detected in a study of the circulating proteome quantified by aptamer-based Somalogic technology^18^. Future studies using mass spectrometry-based absolute levels of urine proteins will provide additional insights.

In conclusion, this first large-scale genetic study of the human proteome in urine shows that the levels of hundreds of highly detectable proteins in urine are influenced by human genetic variation. The results represent a comprehensive genetic framework for understanding kidney-specific processes such as tubular protein reuptake not contained in plasma proteomics studies, highlight potential urine biomarkers of genetic susceptibility to infections and complex diseases, and constitute a resource for mechanistic studies and integration with a wide variety of genetic association studies.

## Online Methods

### Study design and participants

The GCKD study is an ongoing prospective observational cohort study, which recruited 5,217 adult participants with chronic kidney disease (CKD) between 2010 and 2012. Inclusion criteria were an estimated glomerular filtration rate (eGFR) between 30–60 ml/min/1.73m^2^ or an eGFR of >60 with a urinary albumin-to-creatinine ratio (UACR) >300 mg/g or a urinary protein–creatinine ratio of >500 mg/g. The GCKD study was registered in the national registry for clinical studies (DRKS 00003971) and approved by local ethic committees of the participating institutions (universities or medical faculties of Aachen, Berlin, Erlangen, Freiburg, Hannover, Heidelberg, Jena, München and Würzburg)^42^. All participants provided written informed consent. Biomaterials including DNA and urine were collected and were shipped frozen to a central biobank^43^. A more detailed description of the study design, standard operating procedures, and the recruited study population has been published^42,44^. For this project, proteins were quantified from stored spot urine specimens of 1,274 participants with eGFR of >=45 and <=60 ml/min/1.73m^2^ and UACR <300 mg/g at study baseline.

The UK Biobank is a large-scale biomedical research resource containing genetic and health information from half a million UK participants aged 40-69 years at recruitment (2006-2010)^31^. The study received ethical approval from the North West Multi-centre Research Ethics Committee. For this study, we utilized genetic and proteomic data from the UK Biobank Pharma Proteomics Project (UKB-PPP)^20^, which measured 2,923 proteins using the same Olink proximity extension assay technology. Individual-level genetic and proteomic data analyses were performed on the UK Biobank Research Analysis Platform under application ID 64806. Detailed methods for colocalization analyses using summary statistics and individual-level data are described in the “Comparative analyses with plasma pQTLs“ section below.

### Genotyping and imputation

The process of DNA extraction, genotyping and data cleaning in the GCKD study has been described in earlier studies^42,43,45^. Briefly, genomic DNA from GCKD participants was genotyped at 2,612,357 genetic variants using Illumina Omni2.5Exome BeadChip arrays. The quality control steps for the array included checks on individual call rates, sex checks, assessment of heterozygosity, detection of cryptic relatedness, and evaluation of genetic ancestry. At the variant level, SNPs with a call rate below 96% or those failing Hardy-Weinberg equilibrium (HWE) criteria (P<1 × 10⁻⁵) were excluded. After cleaning, the dataset comprised 5,034 individuals. The cleaned genotypes were lifted to build hg38 before imputation. Imputation of the genotypes was performed at the Michigan Imputation Server using Minimac4^46^ using TOPMed Panel r2 as the reference panel and Eagle v2.4 for phasing. Bi-allelic imputed variants with imputation quality of R^2^≥0.3 and a minor allele frequency (MAF) of ≥0.1% were retained, resulting in 15,063,816 high-quality auto- and X-chromosomal variants for GWAS. Only variants with a MAF ≥1% were used for downstream analyses, resulting in a union set of 9,267,849 variants across all tested proteins.

### Proteomic measurement, processing and quality control

All urine samples were centrifugated at 1500g for 10 minutes within 30 minutes of donation as described previously^42,44^. The supernatants were frozen at -80°C in 9 ml tubes and then thawed once for aliquoting into 100 µl vials. The frozen aliquots were shipped to Olink Analysis Service in Sweden (batch 1, N=774), and to Life and Brain Analysis Service in Germany (batch 2, N=500) for proteomic measurements.

Proteins were detected with the Olink Proximity Extension Assay technology^47^, where a matched pair of antibodies labeled with unique complementary oligonucleotides bind to the respective protein target in a sample. The oligonucleotide probes come into close proximity after antibody-binding, hybridize and can be amplified. Protein abundancies are then quantified in a next generation sequencing device. This study used the Olink Explore 3072 library, which targets 2,926 unique proteins grouped into eight different panels (Inflammation I & II, Oncology I & II, Cardiometabolic I & II and Neurological I & II). All proteins are described in **Supplementary Table 2**. The Proximity Extension Assay was first optimized for performance in urine in a pilot study consisting of a dilution series of 16 urine samples. A dilution of 1:1 achieved the best detection results across all protein panels, and our study therefore used undiluted urine samples for the immune reaction of the Proximity Extension Assay.

Data processing of quantified protein levels (NPX values) included the removal of 58 proteins for which 100% of measurements had Olink quality control (QC) warnings, leaving 2,868 proteins for further analysis (**Supplementary Figure 1**). As part of Olink’s built-in QC system, six of the remaining proteins (IL-6, IL-8 (CXCL8), TNF, IDO1, LMOD1 and SCRIB) were detected with four different assays. These were processed separately and the total number of assays (Olink IDs) tested for genetic associations was therefore 2,886.

Prior to genetic analyses, protein levels were corrected for inter-individual variation in urine dilution based on data from 593 proteins (for batch 1) and 659 proteins (for batch 2) measured with <20% of values below the limit of detection using the probabilistic quotient method^21^. Dilution-corrected protein levels were subjected to further QC, resulting in the removal of five samples because they represented outliers (>5 SD) along the first 10 principal components based on proteins with <20% of values below the limit of detection. One sample was excluded because more than 50% of protein measurements hat QC warnings.

### Genome-wide association study of urinary protein levels

Protein level data were intersected with quality-controlled genotype data, resulting in a final dataset of 1,246 (754 batch 1, 492 batch 2, **Supplementary Figure 1**) samples for GWAS. Prior to GWAS, linear regression analyses of protein levels were used to generate residuals adjusted for age, sex, log(eGFR), log(UACR) and the first three genetic principal components, followed by an inverse normal transformation of residuals as the input for GWAS. GWAS of imputed genotype dosages were performed separately in batch 1 and batch 2 as described previously^45^ using SNPTEST v2.5.2 under an additive genetic model. Summary statistics from GWAS in both batches were subjected separately to QC using GWAtoolbox^48^. Subsequently, results from both batches were meta-analyzed using a fixed effects inverse variance model as implemented in METAL (25 March 2011 release 11^49^).

Statistical significance was defined as Bonferroni-corrected genome-wide significance (P <5E-08/2886) in the meta-analysis. Significantly associated SNPs were assigned to loci by selecting, for each protein, the SNP with the lowest genome-wide P-value as the index SNP, by defining the corresponding locus as a 1 Mb interval centered on the index SNP, and by repeating the procedure until no further genome-wide significant SNP remained. For each protein, overlapping intervals were combined, and the variant with the lowest P-value was designated as pQTL. The extended major histocompatibility complex (MHC) region (chromosome 6, 25.5–34 Mb) was considered as one region. Circular plots of association results were created using Circos v0.69-6^50^.

### Variant annotation and gene prioritization

#### Variant annotation

Functional annotation of variants was carried out using VEP v112^51^. We performed VEP annotation for three sets of variants: pQTLs (index variants), credible sets variants with a conditional P-value<5E-8 and filtered for PIP>0.01 outside the MHC region, and the background variant set including all meta-analyzed variants with MAF>0.01. We ran VEP with the options “--canonical --biotype --mane –pick“. In order to have a complete annotation of index variants, we repeated the VEP run without the “--pick“ option.

For the detailed annotation of pQTLs in **Supplementary Table 3** and CS variants in **Supplementary Tables 7 and 8**, we ran VEP with the “—everything“ option, and selected the most severe variant consequences of the MANE transcript encoding the pQTL-associated protein and on any other transcript. Variant overlaps with promoter, enhancer, and open chromatin regions, CTCF binding sites, and transcription factor motifs were also processed and added from the VEP output. The overlap of pQTLs and CS variants with accessible chromatin regions was performed using called peaks in BED format from bulk ATAC-seq data from kidney cortex and medulla^52^ and single-cell ATAC-seq data from various kidney cell types (a Seurat object with called peaks was kindly provided by the authors of the study upon request)^53^. For overlap with candidate *cis*-regulatory regions (ENCODE cCREs v3), a BED file with cCREs was retrieved from https://screen.encodeproject.org/^54^.

#### Gene prioritization

Gene prioritization at each locus was performed by assigning the gene encoding for the respective protein at *cis*-pQTLs. For *trans*-pQTLs, we used an in-house pipeline based on the ProGeM framework^55^. In addition to the bottom-up part of ProGeM, we performed colocalization analyses with transcript levels from GTEx^33^ and kidney gene expression datasets as well as with plasma protein levels^18,20^. At each urine association signal, all protein-coding genes overlapping the 1-Mb region centered on the index SNP were considered. For each gene, the following information was generated and collected: proximity to the index SNP, with the first three closest genes included in downstream ranking; overlap of the gene with the index SNP or good (R^2^>0.8) proxy SNPs (“LD_overlap”); whether the index or proxy SNPs were significant eQTLs of the gene in any of the GTEx v8 tissues (“cis-eQTL”); whether the index or proxy SNPs within a gene were annotated as moderate or high impact in VEP (“IMPACT_moderate_high”); whether the locus colocalized with gene expression in *cis* (“coloc_eQTL”); and whether the locus colocalized with circulating levels of proteins in *cis* (“coloc_pQTL”). Each of these criteria received a score of one, followed by summation. The gene with the highest summary score was assigned as the prioritized gene at *trans*-pQTL. In case of ties, genes were prioritized based on the order coloc_pQTL > coloc_eQTL > IMPACT_moderate_high > *cis*-eQTL > LD_overlap > nearest > 2nd nearest > 3rd nearest gene.

### GO, KEGG, tissue and cell type enrichment analyses

Pathway as well as tissue and cell type enrichment analyses of prioritized genes for **Supplementary Table 4 and 5** were performed by first constructing a database of all Entrez Gene identifiers based on the R package org.Hs.eg.db v3.12.0. The number of independent SNPs per gene was computed using the imputed GCKD genotypes by PLINK version 1.90^56^ with the option “--indep-pairwise 50kb 5 0.2“ and stored in a database. Other stored annotations were gene length, GO terms^57^, KEGG pathways^58^, whether the encoded protein was tissue- or group-enriched or whether it was cell type-enhanced, - enriched or -group enriched in the Human Protein Atlas^35^; whether a gene was among the top 10% highly expressed genes in each tissue of GTEx version 8^33^ or in individual cell types of the human^59–61^ or murine kidney^62^. We then performed 100 million random draws of an equal number of genes as contained in the input list of prioritized genes, matching for deciles of the number of independent SNPs and deciles of gene length, and compared any overlap with cell types, tissues and terms with the ones identified for the original input gene list to obtain empirical p-values. The same analyses were repeated using a database that included only genes encoding for proteins that passed the Olink QC procedure to account for the fact that only approximately 3,000 of 20,000 proteins encoded in the genome were assayed with the Olink technology. Multiple-testing correction was applied using the Benjamini–Hochberg procedure.

For the tissue enrichments based on Human Protein Atlas (HPA) RNA expression data^35^ (**Supplementary Tables 6** and **Figure 3a**), prioritized genes were assigned to a tissue if the respective gene was tissue-enriched, group-enriched or tissue-enhanced for this tissue. Each tissue-gene assignment was then further refined by appending the secretory status (e.g., “secreted to blood“ or “intracellular and membrane“) of a gene to the respective tissue based on the HPA secretome database^34^. For tissue enrichments based on immunohistochemistry (IHC, **Supplementary Table 6** and **Figure 3b**), prioritized genes were assigned to tissues and cellular compartments based on IHC expression grades from the Human Protein Atlas^35^ (≥low: low, medium or high intensity; ≥medium: medium or high intensity; ≥high: high intensity). All enrichment p-values were computed using Fisher’s exact test.

Based on the functional annotation of the 171 unique pQTLs and 1,943 unique credible set variants with a conditional P-value<5E-8 and PIP>0.01 (see “Variant annotation” above), we tested the enrichment of the different annotation categories by Fisher’s exact test compared to the background variant set. Frameshift and stop-gained variants were grouped into one consequence category. For the category “Impact”, we grouped high and moderate into one category. All enrichment results are summarized in **Supplementary Table 9**, while results for annotation categories with more than two annotated index variants are also shown in **Figure 3c**.

### Statistical fine mapping and independent SNP selection

We used sum of single-effects regression (SuSiE, v.0.12.35)^63^ to identify and fine-map independent signals using individual-level genotypes and protein-level measurements from all included participants. Mean-centered and unit-variance additive genotype dosages were used as input, along with batch-corrected, inverse normal-transformed phenotype residuals accounting for the same covariates as in the marginal association colocalization analysis. For each pQTL region, we applied SuSiE regression using the initial parameters: min_abs_corr = 0.1, L = 10, max_iter = 1000, and coverage = 0.95. No region exceeded eight independent credible sets (CS).

Conditional association statistics for all CS variants were computed between additive genotype dosages and inverse normal-transformed residuals (batch-corrected, accounting for covariates from the main analysis) by conditioning each variant on the variant with the highest posterior inclusion probability [PIP] from each additional CS in the region. A collinearity threshold of 0.9 was applied, and the conditional p-value, beta, and standard error were set to 1, 0, and infinity, respectively, if this threshold was exceeded.

### Pairwise colocalization tests

Colocalization analyses were performed using the “coloc” R package^64^ to investigate whether the urine pQTL regions shared underlying genetic variants with other traits. We applied the enumeration approach implemented in the “coloc.abf“ function, considering posterior probabilities for five tested hypotheses (H): both traits have no causal variant in a region (hypothesis H0), either the first or second trait has a causal variant (H1 and H2, respectively), both traits have causal variants and they are distinct (H3), or both traits share a common causal variant (H4). For each significant urine pQTL, we extracted a respective 1-Mb genomic region from the association summary statistics of the respective tested trait for colocalization analysis. Colocalization analyses were only performed if the tested trait contained at least one variant with association P-value <1E-05 in this region. We considered a pair of traits with posterior probability (PP) of H4 >0.8 as colocalized.

In order to rapidly test the 174 urine pQTL regions for shared genetic determinants with thousands of molecular and clinical traits, we set up an in-house colocalization pipeline. Molecular datasets comprised gene expression based on the GTEx Portal (version 8) including 49 tissues and 39,867 unique transcripts^33^, a human kidney eQTL atlas with 14,694 unique transcripts^45^, and two plasma proteomics datasets^18,20^. The plasma protein datasets comprised 2,940 proteins quantified in the UK Biobank Pharma Proteomics using the same technology as the urine proteomics data we generated^20^, as well as 4,909 proteins quantified using complementary, aptamer-based technology from an Icelandic study^18^. Lastly, pairwise colocalization was also performed among the urine pQTL regions. Colocalization analyses between each urine protein and clinical outcomes and diseases were carried out using summary statistics from GWAS of 1419 clinical outcomes in the UK Biobank (downloaded from https://pheweb.org/UKB-TOPMed/) and of 2,272 clinical outcomes that were part of FinnGen release 9^65^. We also performed a targeted investigation of the *PSCA* locus in GWAS meta-analysis for UTI involving 213,869 cases and 1,646,967 controls across several biobanks (UKBB, FinnGen, All-of-Us, MVP, eMERGE, BioVU and 23andMe).

To comprehensively assess whether genetic variants influencing urine protein levels also affect circulating plasma levels of the same protein while accounting for the potential presence of several independently associated variants in a locus, we performed statistical fine-mapping followed by colocalization analysis using individual-level data from the UK Biobank. For the 163 urine pQTL regions outside the MHC, we extracted genotype data from the UK Biobank TOPMed-imputed dataset for European ancestry participants (N=34,090) within 1-Mb windows centered on each urine pQTL. Plasma protein levels were pre-processed by adjusting for age, sex, age*sex interaction, and the first 20 genetic principal components, followed by inverse normal transformation of residuals. We applied SuSiE^63^ for fine-mapping with the following parameters: L=10 credible sets maximum, minimum absolute correlation of 0.1, and using genotypes scaled to unit variance.

Following fine-mapping in both urine (GCKD) and plasma (UKB) datasets, we performed colocalization using the coloc.susie function from the coloc R package^64^. We considered a posterior probability of a shared causal variant (PP.H4) ≥0.8 as evidence for colocalization between a urine and plasma protein signal. For loci where SuSiE identified multiple credible sets, we evaluated colocalization for each set independently and classified a locus as showing shared genetic architecture if any credible set pair showed PP.H4≥0.8.

### Pleiotropy screen

For this screen, statistical significance was defined as genome-wide significance (P<5E-8) in the meta-analysis. Significantly associated SNPs were assigned to pQTLs using the same approach as in the main analysis (see above). Pairwise colocalization tests for pQTLs outside the extended MHC region were performed as described above. To summarize colocalization results, we identified “lead pQTLs“ using a hierarchical ranking system. Within each genomic region, all pQTLs were ranked by the ascending P-value. A pQTL was designated as a lead pQTL if no stronger, colocalized signal existed in the same region. For each lead pQTL, we compiled a comprehensive set of colocalized proteins by combining: (a) proteins directly colocalized with the lead pQTL, and (b) proteins colocalized with any pQTLs assigned to that lead pQTL (reported as Protein.coloc.union in **Supplementary Table 13**).

### Webserver

We developed an interactive web resource with four specialized interfaces for urine pGWAS data exploration: (1) Matrix interface: Interactive heatmap visualization displaying protein-loci associations with detailed table views for filtering associations by loci, protein targets, p-value thresholds and effect sizes. The underlying data, structured in JSON format, is processed and rendered using a custom based matrix builder based on D3.js^66^ (version 4.12.2). (2) Network interface: Visualization showing colocalization and association relationships with interactive locus and distance selection, detailed tooltips, and dynamic filtering by p-value and node types. The complete network was prepared using Cytoscape^67^ (version 3.10.3) desktop software, then exported and subsequently imported into the web resource through Cytoscape.js^68^. (3) Table interface: Searchable data table with column sorting and filtering by genomic coordinates, gene symbols or protein targets. Table processing and interface are provided by the DataTables (version 1.10.20, http://www.datatables.net) library. Data can be exported to clipboard or comma separated values file. (4) Regional association plot interface: Regional association plots integrating gene annotations and recombination rate data visualization based on dropdown selections. Tabix^69^-compressed result files are rendered by LocusZoom^70^ (version 0.14.0) using built-in and custom data parsing. For building the web resource, we utilized the javascript libraries: jQuery (version 2.2.0, https://jquery.com), jQueryUI (version 1.12.1, https://jqueryui.com) and the Strata website template by HTML5 UP (https://html5up.net).

### Generative AI language models

We used generative AI language models (Claude Opus 4, Anthropic) throughout the manuscript preparation process for iterative drafting and refinement of content. All AI-generated contributions were reviewed, verified, and edited by the authors. We take full responsibility for the accuracy, integrity, and originality of all content in the final manuscript.

## Data availability

Summary statistics for all significant gene–protein associations (pQTLs and credible sets) are available in **Supplementary Tables 3, 7, and 8**. The results from genetic colocalization analyses with molecular and clinical traits are provided in **Supplementary Tables 10-15**.

All proteo-genomic results and genome-wide summary association data are accessible through the following interactive portal: http://gwas.eu/urine-p/ **(freely available after successful peer-reviewed publication**) and through the NHGRI-EBI GWAS Catalog (ID will be added after publication, https://www.ebi.ac.uk/gwas/).

Detailed information on used external datasets is provided in the respective sections of the Methods.

## Code availability

For general coding, R (versions 3.6.3 and 4.0.5) was used. Detailed information on used software and options is provided in the respective sections of the Methods.

## Acknowledgements

The work of S.H., O.B., N.S., M.W., and A.K. was funded by German Research Foundation (DFG) project ID 431984000 (SFB 1453). The work of N.S. and A.K. was funded by DFG project ID 499552394 (SFB 1597/1). Germany’s Excellence Strategy (CIBSS, EXC-2189, project ID 390939984) supported the work of Y.L. and A.K. The work of A.K. was also supported by DFG project ID 441891347 (SFB-1479). The work of K.X. is supported by National Institute of Diabetes and Digestive and Kidney Diseases (NIDDK) grant K01-DK135917. A.K. is supported by NIDDK grants R03-DK144285 and K25-DK128563. K.K. is supported by NIDDK grants R01-DK144793, R01-DK105124, R01-DK136765, and National Human Genome Research Institute (NHGRI) grants U01-HG013201 and U01-HG008680.

Genotyping in the GCKD study was supported by Bayer Pharma. The GCKD study was and is supported by the BMBF (FKZ 01ER 0804, 01ER 0818, 01ER 0819, 01ER 0820 and 01ER 0821) and the KfH Foundation for Preventive Medicine. Unregistered grants to support the study were provided by corporate sponsors (listed at https://gckd.org). We are grateful for the willingness of the patients to participate in the GCKD study. The enormous effort of the study personnel of the various regional centers is highly appreciated. We thank the large number of nephrologists who provide routine care for the patients and collaborate with the GCKD study. The GCKD investigators are listed in the Supplementary Note.

## Author Contributions

Design of this study: A.K.

Recruitment and management of study: A.K., S.H.

Processing of genetic data: M.W., Y.L.

Bioinformatics and statistical analysis: A.K., S.H., I.M., O.B., Y.L., B.U., N.S., P.Sc.

Webserver: N.L., W.R.M., G.K., S.H., M.S., M.A.

Interpretation of Results: A.K., S.H., O.B., Y.L.

External data contributions: K.X., A.Kh., K.K.

Wrote the manuscript: A.K., S.H., O.B., Y.L.

Critically read and approved the manuscript: S.H., O.B., M.S., I.M., K.X., A.Kh., N.S., S.M.M, B.U., M.A., P.Sc., P.Se, F.K., K.-U.E., K.K., M.W. Y.L., A.K.

## Conflicts of interest

The authors report no conflict of interest.

## Notes

### Competing Interest Statement

The authors have declared no competing interest.

### Author Declarations

Ethics Committees of all participating study centers gave ethical approval for the GCKD study wich provided all patient samples and data for this study. German Clinical Trials Register number is DRKS 00003971. The study centers are: Friedrich-Alexander-University Erlangen-Nuernberg, Medical Faculty of the Rheinisch-Westfaelische Technische Hochschule Aachen, Charite University Medicine Berlin, Medical Center University of Freiburg, Medizinische Hochschule Hannover, Medical Faculty of the University of Heidelberg, Friedrich-Schiller-University Jena, Medical Faculty of the Ludwig-Maximilians-University Munich, Medical Faculty of the University of Wuerzburg.

## References

1. Zhao, M. et al. A comprehensive analysis and annotation of human normal urinary proteome. Sci Rep 7, 3024 (2017).

2. Maack, T., Johnson, V., Kau, S. T., Figueiredo, J. & Sigulem, D. Renal filtration, transport, and metabolism of low-molecular-weight proteins: A review. Kidney International 16, 251–270 (1979).

3. Adachi, J., Kumar, C., Zhang, Y., Olsen, J. V. & Mann, M. The human urinary proteome contains more than 1500 proteins, including a large proportion of membrane proteins. Genome Biol 7, R80 (2006).

4. Matsushita, K. et al. Estimated glomerular filtration rate and albuminuria for prediction of cardiovascular outcomes: a collaborative meta-analysis of individual participant data. Lancet Diabetes Endocrinol 3, 514–525 (2015).

5. Nielsen, R., Christensen, E. I. & Birn, H. Megalin and cubilin in proximal tubule protein reabsorption: from experimental models to human disease. Kidney International 89, 58–67 (2016).

6. Gräsbeck, R. Imerslund-Gräsbeck syndrome (selective vitamin B12 malabsorption with proteinuria). Orphanet J Rare Dis 1, 17 (2006).

7. Kantarci, S. et al. Mutations in LRP2, which encodes the multiligand receptor megalin, cause Donnai-Barrow and facio-oculo-acoustico-renal syndromes. Nat Genet 39, 957–959 (2007).

8. Aminoff, M. et al. Mutations in CUBN, encoding the intrinsic factor-vitamin B12 receptor, cubilin, cause hereditary megaloblastic anaemia 1. Nat Genet 21, 309–313 (1999).

9. Weiss, G. L. et al. Architecture and function of human uromodulin filaments in urinary tract infections. Science 369, 1005–1010 (2020).

10. Devuyst, O., Olinger, E. & Rampoldi, L. Uromodulin: from physiology to rare and complex kidney disorders. Nat Rev Nephrol 13, 525–544 (2017).

11. Fassett, R. G. et al. Biomarkers in chronic kidney disease: a review. Kidney International 80, 806–821 (2011).

12. Carrasco-Zanini, J. et al. Mapping biological influences on the human plasma proteome beyond the genome. Nat Metab 6, 2010–2023 (2024).

13. Deng, Y.-T. et al. Atlas of the plasma proteome in health and disease in 53,026 adults. Cell 188, 253–271.e7 (2025).

14. Suhre, K. et al. Connecting genetic risk to disease end points through the human blood plasma proteome. Nat Commun 8, 14357 (2017).

15. Emilsson, V. et al. Co-regulatory networks of human serum proteins link genetics to disease. Science 361, 769–773 (2018).

16. Sun, B. B. et al. Genomic atlas of the human plasma proteome. Nature 558, 73–79 (2018).

17. Pietzner, M. et al. Mapping the proteo-genomic convergence of human diseases. Science 374, eabj1541 (2021).

18. Ferkingstad, E. et al. Large-scale integration of the plasma proteome with genetics and disease. Nat Genet 53, 1712–1721 (2021).

19. Gudjonsson, A. et al. A genome-wide association study of serum proteins reveals shared loci with common diseases. Nat Commun 13, 480 (2022).

20. Sun, B. B. et al. Plasma proteomic associations with genetics and health in the UK Biobank. Nature 622, 329–338 (2023).

21. Schlosser, P. et al. Genetic studies of paired metabolomes reveal enzymatic and transport processes at the interface of plasma and urine. Nat Genet 55, 995–1008 (2023).

22. Eckardt, K.-U. et al. The German Chronic Kidney Disease (GCKD) study: design and methods. Nephrol Dial Transplant 27, 1454–1460 (2012).

23. Hussey, A. J. & Hayes, J. D. Characterization of a human class-Theta glutathione S-transferase with activity towards 1-menaphthyl sulphate. Biochem J 286 **(Pt** **3**), 929–935 (1992).

24. Witecka, A. et al. Hydroxysteroid 17-β dehydrogenase 14 (HSD17B14) is an L-fucose dehydrogenase, the initial enzyme of the L-fucose degradation pathway. J Biol Chem 300, 107501 (2024).

25. Christensen, E. I., Birn, H., Storm, T., Weyer, K. & Nielsen, R. Endocytic receptors in the renal proximal tubule. Physiology (Bethesda) 27, 223–236 (2012).

26. Eshbach, M. L. & Weisz, O. A. Receptor-Mediated Endocytosis in the Proximal Tubule. Annu Rev Physiol 79, 425–448 (2017).

27. Hall, A. M. Protein handling in kidney tubules. Nat Rev Nephrol 21, 241–252 (2025).

28. Storm, T. et al. A patient with cubilin deficiency. N Engl J Med 364, 89–91 (2011).

29. Burmeister, R. et al. A two-receptor pathway for catabolism of Clara cell secretory protein in the kidney. J Biol Chem 276, 13295–13301 (2001).

30. Tzur, S., Wasser, W. G., Rosset, S. & Skorecki, K. Linkage disequilibrium analysis reveals an albuminuria risk haplotype containing three missense mutations in the cubilin gene with striking differences among European and African ancestry populations. BMC Nephrol 13, 142 (2012).

31. Bycroft, C. et al. The UK Biobank resource with deep phenotyping and genomic data. Nature 562, 203–209 (2018).

32. Zhao, B., Tu, C., Shen, S., Qu, J. & Morris, M. E. Identification of Potential Megalin/Cubilin Substrates Using Extensive Proteomics Quantification from Kidney Megalin-Knockdown Mice. AAPS J 24, 109 (2022).

33. GTEx Consortium. The GTEx Consortium atlas of genetic regulatory effects across human tissues. Science 369, 1318–1330 (2020).

34. Uhlén, M. et al. The human secretome. Sci Signal 12, eaaz0274 (2019).

35. Uhlén, M. et al. Proteomics. Tissue-based map of the human proteome. Science 347, 1260419 (2015).

36. Chen, J., Epstein, M. P., Schildkraut, J. M. & Kar, S. P. Mapping inherited genetic variation with opposite effects on autoimmune disease and cancer identifies candidate drug targets associated with the anti-tumor immune response. medRxiv 2023.12.23.23300491 (2023) doi:10.1101/2023.12.23.23300491.

37. Western, D. et al. Proteogenomic analysis of human cerebrospinal fluid identifies neurologically relevant regulation and implicates causal proteins for Alzheimer’s disease. Nat Genet 56, 2672–2684 (2024).

38. GCKD Investigators et al. Genetic studies of urinary metabolites illuminate mechanisms of detoxification and excretion in humans. Nat Genet 52, 167–176 (2020).

39. Scherer, N. et al. Coupling metabolomics and exome sequencing reveals graded effects of rare damaging heterozygous variants on gene function and human traits. Nat Genet 57, 193–205 (2025).

40. Katz, D. H. et al. Proteomic profiling platforms head to head: Leveraging genetics and clinical traits to compare aptamer- and antibody-based methods. Sci Adv 8, eabm5164 (2022).

41. Schlosser, P. et al. Genetic studies of paired metabolomes reveal enzymatic and transport processes at the interface of plasma and urine. Nat Genet 10.1038/s41588-023-01409-8 (2023) doi:10.1038/s41588-023-01409-8.

42. Eckardt, K.-U. et al. The German Chronic Kidney Disease (GCKD) study: design and methods. Nephrology Dialysis Transplantation 27, 1454–1460 (2012).

43. Prokosch, H.-U. et al. Designing and implementing a biobanking IT framework for multiple research scenarios. Stud Health Technol Inform 180, 559–563 (2012).

44. Titze, S. et al. Disease burden and risk profile in referred patients with moderate chronic kidney disease: composition of the German Chronic Kidney Disease (GCKD) cohort. Nephrology Dialysis Transplantation 30, 441–451 (2015).

45. Li, Y. et al. Genome-Wide Association Studies of Metabolites in Patients with CKD Identify Multiple Loci and Illuminate Tubular Transport Mechanisms. J Am Soc Nephrol 29, 1513–1524 (2018).

46. Das, S. et al. Next-generation genotype imputation service and methods. Nat Genet 48, 1284–1287 (2016).

47. Wik, L. et al. Proximity Extension Assay in Combination with Next-Generation Sequencing for High-throughput Proteome-wide Analysis. Mol Cell Proteomics 20, 100168 (2021).

48. Fuchsberger, C., Taliun, D., Pramstaller, P. P., Pattaro, C., & CKDGen consortium. GWAtoolbox: an R package for fast quality control and handling of genome-wide association studies meta-analysis data. Bioinformatics 28, 444–445 (2012).

49. Willer, C. J., Li, Y. & Abecasis, G. R. METAL: fast and efficient meta-analysis of genomewide association scans. Bioinformatics 26, 2190–2191 (2010).

50. Krzywinski, M. et al. Circos: an information aesthetic for comparative genomics. Genome Res 19, 1639–1645 (2009).

51. McLaren, W. et al. The Ensembl Variant Effect Predictor. Genome Biol 17, 122 (2016).

52. Haug, S. et al. Multi-omic analysis of human kidney tissue identified medulla-specific gene expression patterns. Kidney Int 105, 293–311 (2024).

53. Muto, Y. et al. Single cell transcriptional and chromatin accessibility profiling redefine cellular heterogeneity in the adult human kidney. Nat Commun 12, 2190 (2021).

54. ENCODE Project Consortium et al. Expanded encyclopaedias of DNA elements in the human and mouse genomes. Nature 583, 699–710 (2020).

55. Stacey, D. et al. ProGeM: a framework for the prioritization of candidate causal genes at molecular quantitative trait loci. Nucleic Acids Res 47, e3 (2019).

56. Chang, C. C. et al. Second-generation PLINK: rising to the challenge of larger and richer datasets. Gigascience 4, 7 (2015).

57. Gene Ontology Consortium et al. The Gene Ontology knowledgebase in 2023. Genetics 224, iyad031 (2023).

58. Kanehisa, M., Furumichi, M., Sato, Y., Matsuura, Y. & Ishiguro-Watanabe, M. KEGG: biological systems database as a model of the real world. Nucleic Acids Res 53, D672–D677 (2025).

59. Aizarani, N. et al. A human liver cell atlas reveals heterogeneity and epithelial progenitors. Nature 572, 199–204 (2019).

60. Stewart, B. J. et al. Spatiotemporal immune zonation of the human kidney. Science 365, 1461–1466 (2019).

61. Lake, B. B. et al. An atlas of healthy and injured cell states and niches in the human kidney. Nature 619, 585–594 (2023).

62. Park, J. et al. Single-cell transcriptomics of the mouse kidney reveals potential cellular targets of kidney disease. Science 360, 758–763 (2018).

63. Wang, G., Sarkar, A., Carbonetto, P. & Stephens, M. A Simple New Approach to Variable Selection in Regression, with Application to Genetic Fine Mapping. Journal of the Royal Statistical Society Series B: Statistical Methodology 82, 1273–1300 (2020).

64. Giambartolomei, C. et al. Bayesian test for colocalisation between pairs of genetic association studies using summary statistics. PLoS Genet 10, e1004383 (2014).

65. Kurki, M. I. et al. FinnGen provides genetic insights from a well-phenotyped isolated population. Nature 613, 508–518 (2023).

66. Bostock, M., Ogievetsky, V. & Heer, J. D^3^ Data-Driven Documents. IEEE Trans. Visual. Comput. Graphics 17, 2301–2309 (2011).

67. Shannon, P. et al. Cytoscape: A Software Environment for Integrated Models of Biomolecular Interaction Networks. Genome Res. 13, 2498–2504 (2003).

68. Franz, M. et al. Cytoscape.js: a graph theory library for visualisation and analysis. Bioinformatics 32, 309–311 (2016).

69. Li, H. Tabix: fast retrieval of sequence features from generic TAB-delimited files. Bioinformatics 27, 718–719 (2011).

70. Boughton, A. P. et al. LocusZoom.js: interactive and embeddable visualization of genetic association study results. Bioinformatics 37, 3017–3018 (2021).

