## Supplementary Figures for "Insights into Renal Protein Handling Through GWAS of the Human Urine Proteome"

Content

**Affiliations:** ____________________ Fehler! Textmarke nicht definiert.

***Supplementary Figures*** ____________________ ***3***

**Supplementary Figure 1: Study design workflow showing sample and protein selection** ______ **3**

**Supplementary Figure 2: Distribution of pQTL effect sizes across frequencies of the minor allele** **4**

**Supplementary Figure 3: Colocalization analysis of urine proteins and metabolites identifies HSD17B14 as a fucose dehydrogenase** ____________________ **5**

**Supplementary Figure 4: Regional association plots for urine proteins at the *CUBN* locus** _____ **6**

***References for supplementary material*** ____________________ ***7***

***Supplementary Acknowledgements*** ____________________ ***8***

**List of GCKD Study Investigators** ____________________ **8**

### Supplementary Figures

##### Supplementary Figure 1: Study design workflow showing sample and protein selection

Sample and protein selection workflow. The Olink Explore 3072 technology has the capacity to detect 2,926 distinct proteins, which are measured by 2,944 assays. Six proteins (IL-6, IL-8 [CXCL8], TNF, IDO1, LMOD1, and SCRIB) are each detected by four different assays for quality control purposes, resulting in 18 additional assays beyond the number of distinct proteins.

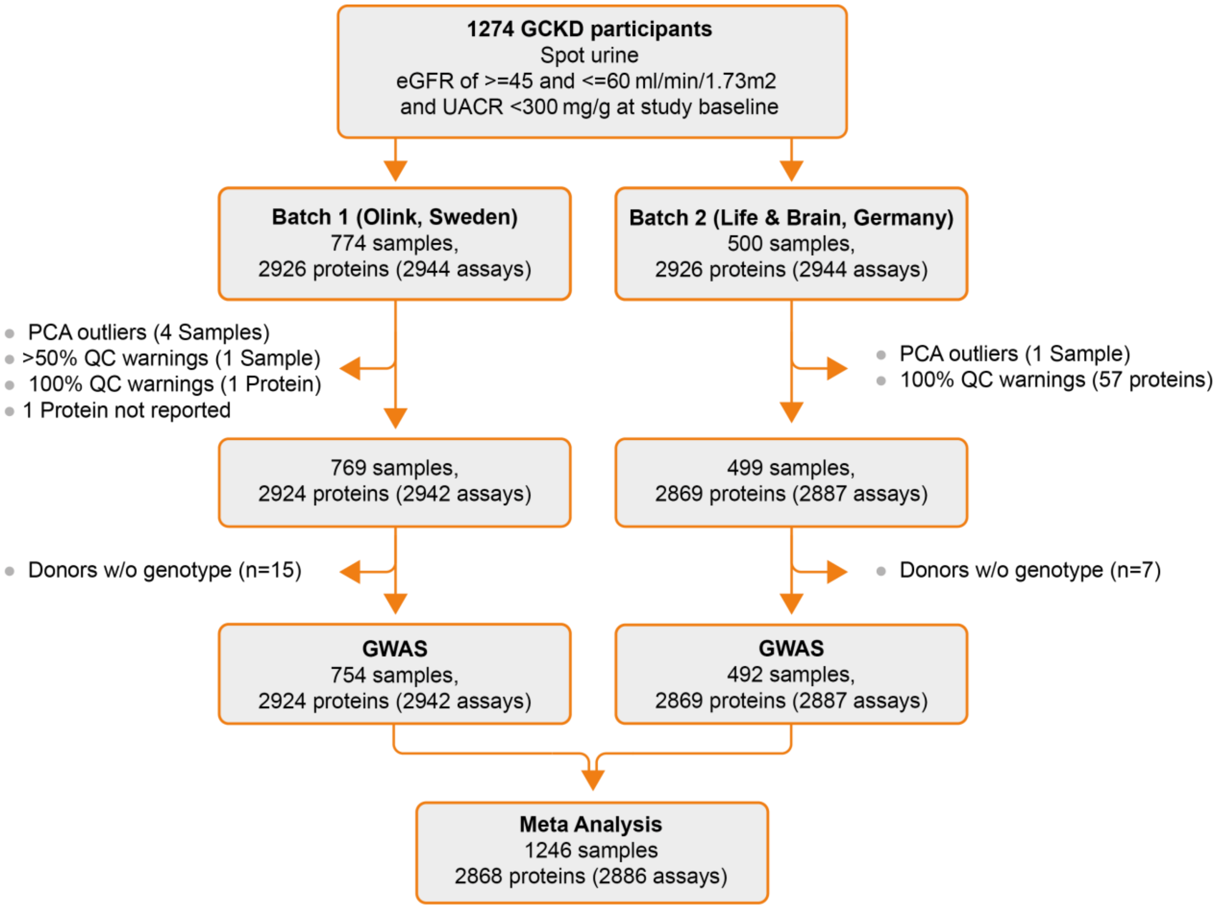

**Supplementary Figure 2: Distribution of pQTL effect sizes across frequencies of the minor allele**

*Cis* and *trans* pQTLs are color-coded as indicated.

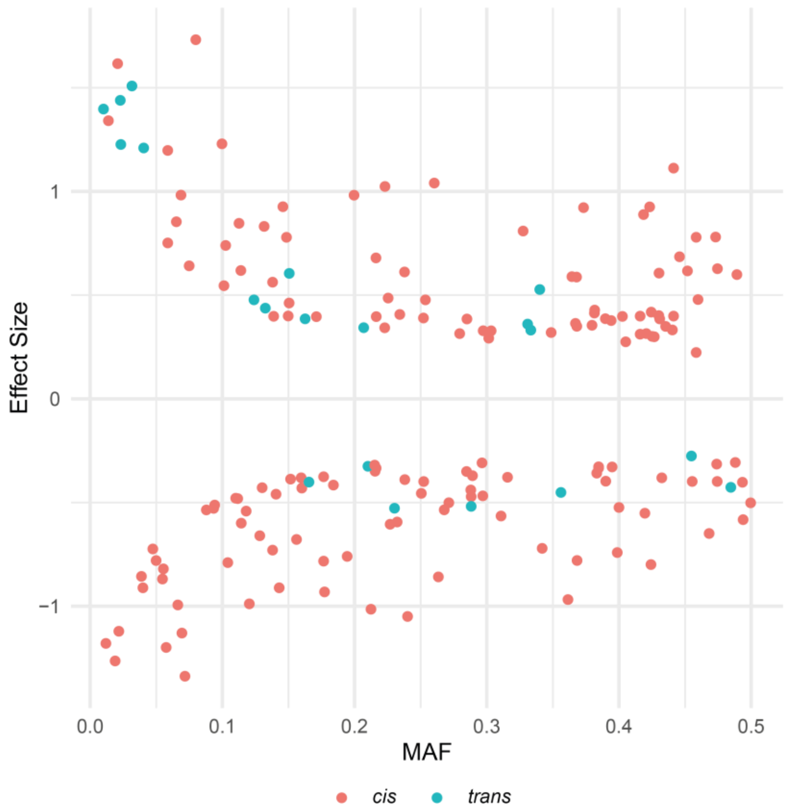

**Supplementary Figure 3: Colocalization analysis of urine proteins and metabolites identifies HSD17B14 as a fucose dehydrogenase**

**(a)** Genetic colocalization of association signals between urine levels of HSD17B14, fucose and arabinose (both colocalize with PP.H4 = 0.98). **(b)** The index variant at the *HSD17B14* locus is a missense variant causing an amino acid substitution from arginine (Arg) to tryptophan (Trp) at position 130. The major G-allele (allele frequency 0.95) encodes arginine and is associated with higher HSD17B14 levels in urine and plasma, and with lower fucose and arabinose levels in urine. HSD17B14 exhibits L-fucose dehydrogenase activity, and both fucose and arabinose are known substrates, with L-fucono-1,5-lactone as the suggested product from fucose dehydrogenation[1]. **(c)** Ribbon representation of human HSD17B14 in complex with NAD (black sticks) and estrone (red sticks) published by Witecka *et al.*[1] (left panel). The right panel highlights the position of arginine 130 (AlphaFold prediction[2]).

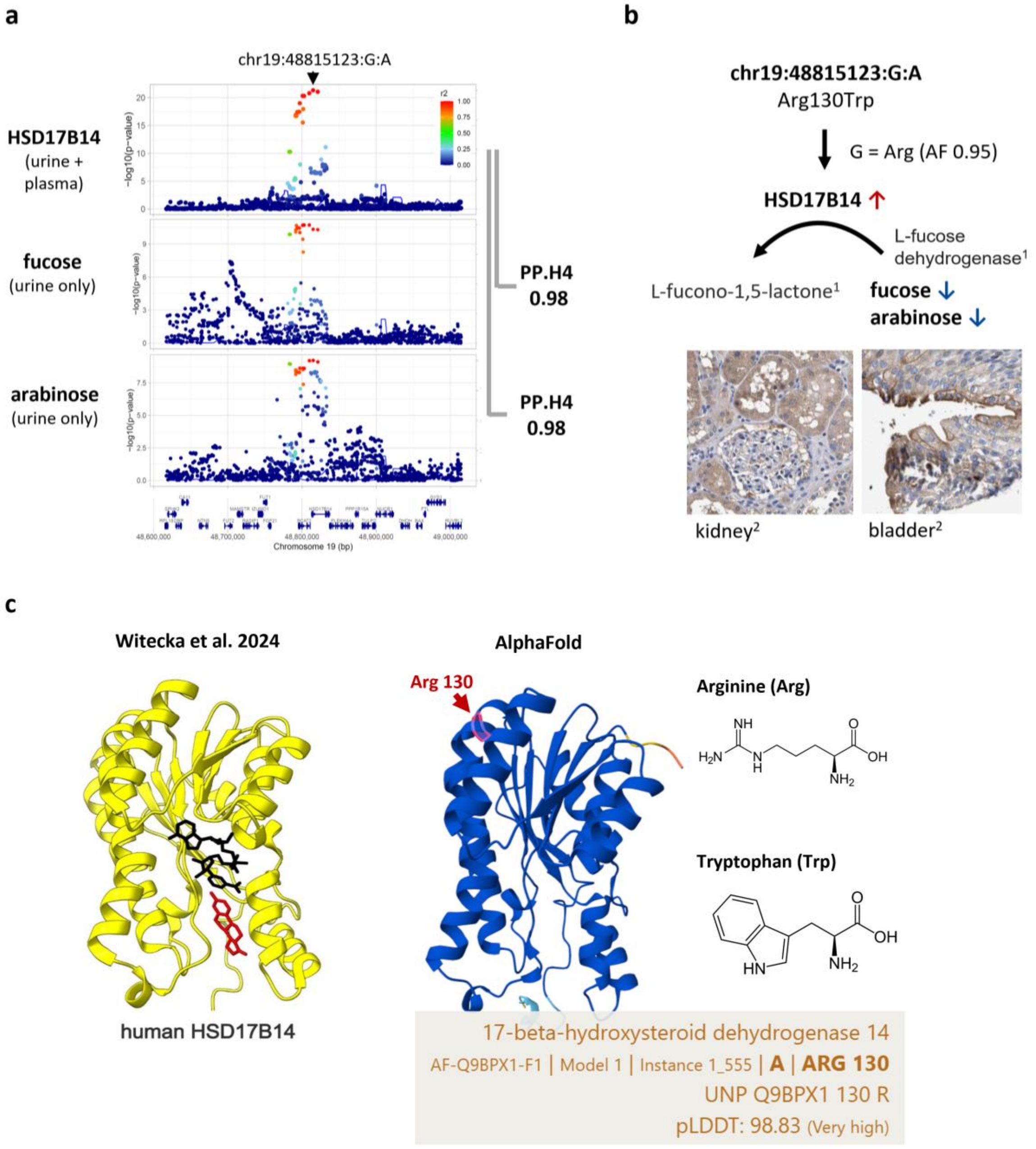

#### Supplementary Figure 4: Regional association plots for urine proteins at the *CUBN* locus

Regional association plots at the *CUBN* locus for urine proteins with association signals that show genetic colocalization with the primary IFNGR1 association (top left panel). “Locus 1” refers to the locus number from Supplementary Table 13 (colocalization-based pleiotropy screen). The displayed proteins are listed in the "Protein.coloc.union" column. Only proteins with a minimum p-value <1E-6 in this locus are shown. $r^2$: linkage disequilibrium with the index variant of the pQTL.

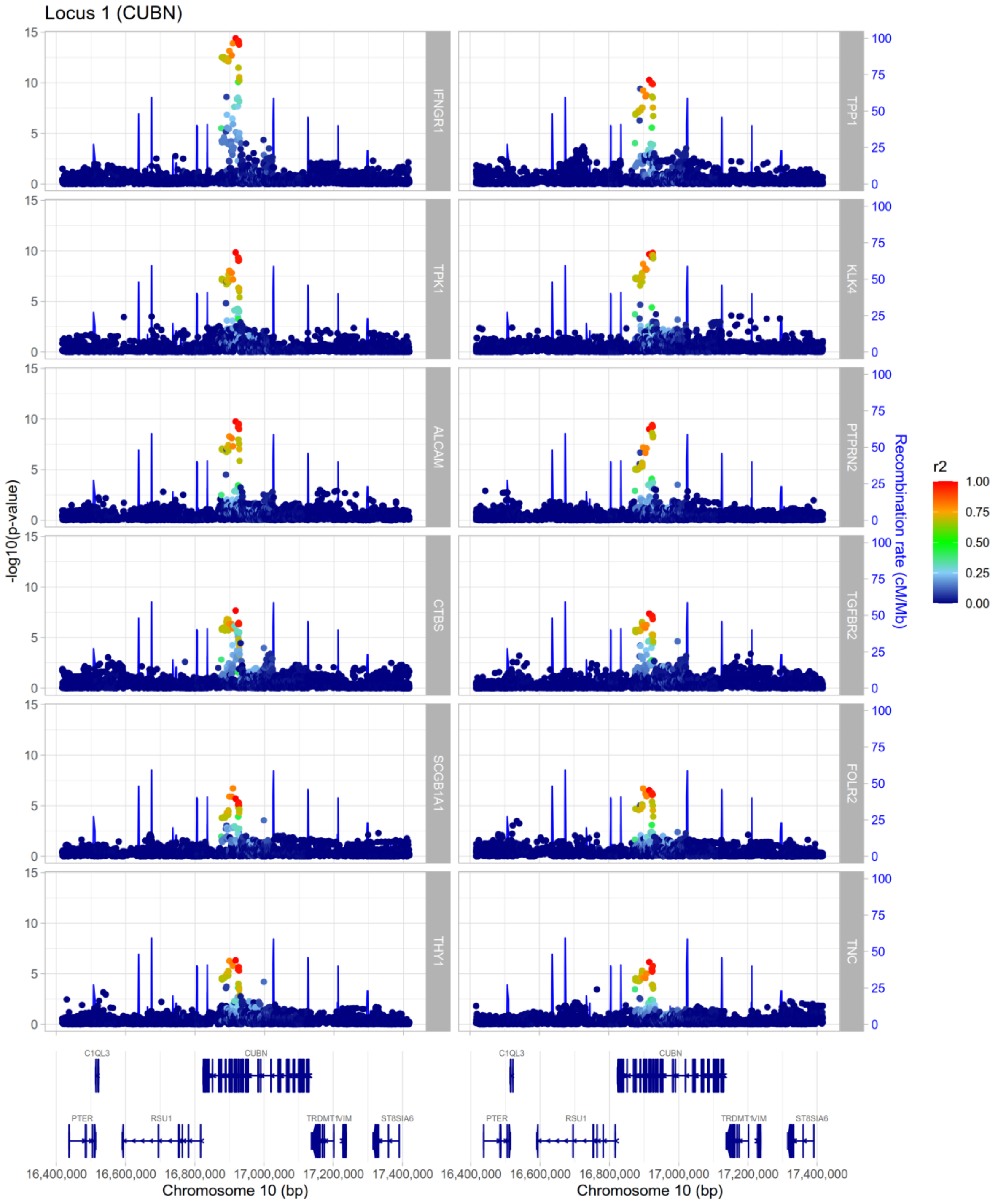

### References for supplementary material

1. Witecka, A. *et al.* Hydroxysteroid 17-β dehydrogenase 14 (HSD17B14) is an L-fucose dehydrogenase, the initial enzyme of the L-fucose degradation pathway. *J Biol Chem* **300**, 107501 (2024).
2. Jumper, J. *et al.* Highly accurate protein structure prediction with AlphaFold. *Nature* **596**, 583–589 (2021).

### Supplementary Acknowledgements

#### List of GCKD Study Investigators

A list of nephrologists currently collaborating with the GCKD study is available at http://www.gckd.org.

| | |
|---|---|
| University of Erlangen-Nürnberg | Kai-Uwe Eckardt, Heike Meiselbach, Markus Schneider, Mario Schiffer, Hans-Ulrich Prokosch, Barbara Bärthlein, Andreas Beck, André Reis, Arif B. Ekici, Susanne Becker, Ulrike Alberth-Schmidt, Anke Weigel, Sabine Marschall |
| University of Freiburg | Gerd Walz, Anna Köttgen, Ulla Schultheiß, Wibke Bechtel-Walz, Fruzsina Kotsis, Simone Meder, Erna Mitsch, Ursula Reinhard |
| RWTH Aachen University | Jürgen Floege, Rafael Kramann, Turgay Saritas |
| Charité, University Medicine Berlin | Elke Schaeffner, Seema Baid-Agrawal, Kerstin Theisen |
| Hannover Medical School | Kai Schmidt-Ott |
| University of Heidelberg | Martin Zeier, Claudia Sommerer |
| University of Jena | Gunter Wolf, Martin Busch, |
| Ludwig-Maximilians University of München | Thomas Sitter |
| University of Würzburg | Christoph Wanner, Vera Krane, Britta Bauer |
| Medical University of Innsbruck, Division of Genetic Epidemiology | Florian Kronenberg, Julia Raschenberger, Barbara Kollerits, Lukas Forer, Sebastian Schönherr, Hansi Weissensteiner |
| University of Regensburg, Institute of Functional Genomics | Peter Oefner, Wolfram Gronwald |
| Department of Medical Biometry, Informatics and Epidemiology (IMBIE), University of Bonn | Matthias Schmid, Jennifer Nadal |
